# Digital self-efficacy as a potential intermediary between vision impairment and daily internet use among older adults: A cross-sectional analysis of HINTS 2024

**DOI:** 10.64898/2026.06.09.26353388

**Authors:** Haruno Suzuki, Thomas Hoffmann, Heather Leutwyler, Margaret Wallhagen

## Abstract

**Background:** Older adults with vision impairment often experience barriers to using digital technology. The indirect associations between vision impairment and digital access and skills via digital self-efficacy and frustration among older adults remain largely unknown.

**Objective:** This study aimed to 1) explore factors associated with digital access, skills, self-efficacy, and frustration among older adults with vision impairment; 2) examine associations between vision impairment and digital access, skills, self-efficacy, and frustration among older adults; and 3) examine whether digital self-efficacy and frustration may help explain associations between vision impairment and digital access and skills among older adults.

**Methods:** This was a cross-sectional study using nationally representative data from the Health Information National Trends Survey (HINTS) 2024. Respondents aged 60 and older were included. Vision impairment was assessed using a self-reported item. Outcomes included self-reported digital access, skills, self-efficacy, and frustration. Survey-weighted multivariable logistic regression and generalized structural equation modeling were conducted, adjusting for age, sex, race/ethnicity, education, and the number of comorbidities.

**Results:** Among 3,149 older adults (mean [SD] age, 70.7 [10.0] years; 45.6% female), 7.1% (n=223) reported vision impairment. Among older adults with vision impairment, 65.6% (95% CI, 53.5% to 75.9%) used the internet daily, and 79.5% (95% CI, 66.8% to 88.2%) used a smartphone in the past 12 months. In multivariable logistic regression analyses among older adults with vision impairment, older age was associated with lower odds of daily internet use (OR, 0.84; 95% CI, 0.79 to 0.90), smartphone use (OR, 0.85; 95% CI, 0.75 to 0.97), wearable device use (OR, 0.88; 95% CI, 0.79 to 0.97), and using the internet to send a message to a healthcare provider (OR, 0.87; 95% CI, 0.80 to 0.93). Older adults who self-identified as racial and ethnic minority groups (e.g., Black/African American, Hispanic) had lower odds of daily internet use (OR, 0.15; 95% CI, 0.05 to 0.50) and using the internet to send a message to a healthcare provider (OR, 0.17; 95% CI, 0.04 to 0.73) compared with Non-Hispanic White older adults. Vision impairment was associated with lower odds of daily internet use (OR, 0.60; 95% CI, 0.37 to 0.99) and digital self-efficacy (OR, 0.53; 95% CI, 0.32 to 0.86). Digital self-efficacy was associated with higher odds of daily internet use (OR, 2.95; 95% CI, 2.04 to 4.26). Generalized structural equation modeling identified an indirect association between vision impairment and daily internet use via digital self-efficacy (coefficient, -0.68; 95% CI, -1.24 to -0.12).

**Conclusions:** Findings suggest that reduced digital self-efficacy may help explain the observed association between vision impairment and daily internet use among older adults. Interventions targeting digital self-efficacy, including accessible interface designs, personalized coaching, and peer support, may help bridge the digital divide among older adults with vision impairment.

## Introduction

The global burden of vision impairment is predicted to escalate rapidly. Approximately 600 million people worldwide had distance vision impairment or blindness in 2020 [1]. Vision impairment and blindness are most common among adults aged 50 or older, largely attributable to age-related eye conditions, such as cataracts, age-related macular degeneration, glaucoma, and diabetic retinopathy [2]. As the global population ages, the prevalence of vision impairment and blindness is projected to reach 895 million by 2050, with a 50% increase from 2020 [1]. This trend is similarly evident in the United States, where more than one-quarter of adults aged 71 or older reported having vision impairment in 2021 [3]. Vision impairment is associated with a wide range of adverse health outcomes, including functional limitations [4], depression [5], cognitive decline and dementia [6], increased mortality [7], and reduced quality of life [8]. Maintaining functional independence and enhancing the quality of life among older adults with vision impairment is therefore a global public health priority.

Digital technology has the potential to improve health outcomes and support independent living among older adults with vision impairment. However, vision impairment presents substantial barriers to accessing and using digital technology in healthcare settings. For example, telehealth is a promising tool for delivering virtual healthcare to diverse older adults without in-person care, particularly for individuals facing transportation challenges to healthcare facilities, including those with mobility limitations and those living in rural areas [9]. However, these online platforms rely heavily on visual information, which poses interface-related challenges for older adults with vision impairment, such as small text, low color contrast, the lack of alternative text for images, websites that are not compatible with screen reader software, and limited audio descriptions during video calls [10,11]. Prior review papers identified key barriers to digital technology access and use among older adults, including limited accessibility of websites and digital devices, low digital health literacy, high technology costs, privacy concerns, and mistrust of digital platforms [10,12–14]. One mixed-method study found that vision-related challenges restrict several internet-based instrumental activities of daily living, including online shopping, transportation via mobile apps, and monitoring health [11]. Therefore, addressing these barriers is urgently needed to ensure equitable access to digital health care for this population.

Digital divide is defined as “inequalities in individuals’ access, skills, and outcomes in using digital technology” [15–17]. Digital divide encompasses three dimensions: the first-level digital divide (digital access divide), refers to inequalities in access to digital devices and broadband connectivity [17,18]; the second-level digital divide (digital skill divide), refers to inequalities in the ability to use the internet and digital devices [17,19]; and the third-level digital divide (digital outcome divide), refers to differences in the benefits derived from digital technology use [17,20]. These divides disproportionately affect socially vulnerable populations. For example, older adults, racial and ethnic minorities, individuals with lower educational attainment, those with lower household income, and those with functional impairment are at high risk of experiencing a digital divide [21–23]. Given that older adults with vision impairment are at the intersection between older age and functional impairment, prior studies reported that older adults with vision impairment have a lower proportion of daily internet use and digital device ownership compared with those without vision impairment [24–26]. Among older adults, older age, racial and ethnic minorities, individuals with lower educational attainment, those with lower household income, and a greater number of comorbidities are associated with lower odds of accessing the internet and having a smartphone or tablet [21]. However, there are limited studies that explored factors associated with low digital access and skills focusing on older adults with vision impairment.

The Senior Technology Acceptance Model (STAM) [27], which embodies Bandura’s social cognitive theory [28], is a useful conceptual framework to understand the unique barriers to using digital technology among older adults. STAM encompasses digital self-efficacy and frustration as key determinants of technology usage behaviors among older adults [27,29]. Digital self-efficacy is defined as an individual’s perception of their capability to perform tasks related to the use of digital technology [28,30]. Digital self-efficacy is associated with higher digital health literacy scores [31] and lower anxiety scores about learning new digital technology [32] among adults. Within STAM, the construct, ‘gerontechnology self-efficacy,’ which is defined as older adults’ sense of being able to use technology successfully [27], is closely related to digital self-efficacy and recognized as a key predictor of technology acceptance and use among older adults. Digital frustration is defined as an emotional response to a failure associated with using digital technology [33]. This construct is a newly emerging concept in digital health research. Digital frustration builds upon computer frustration research, which conceptualizes computer frustration as an emotional response generated by the user’s appraisal of interface performance as an impediment to goal attainment [34]. Technology anxiety, another form of negative emotional response to using digital technology, is associated with lower behavioral intention to use digital health technologies [35–40], avoidance of using digital tools [41], and lower online health information seeking behaviors [42] among older adults. Although digital frustration is conceptually related to the ‘gerontechnology anxiety’ construct in STAM, which is defined as older adults’ apprehension when faced with the possibility of using technology [27], the two constructs differ in their temporal focus: digital frustration captures the emotional response during digital technology use following a failure, whereas gerontechnology anxiety involves anticipatory apprehension before use. Despite their theoretical importance in STAM, whether vision impairment is associated with lower digital self-efficacy and higher digital frustration among older adults, and whether there is an indirect association from vision impairment to digital access and skills via digital self-efficacy and frustration among older adults, remains largely unknown.

To address these knowledge gaps, this study aims to: 1) explore factors associated with digital access, skills, self-efficacy, and frustration among older adults with vision impairment; 2) examine the association between vision impairment and digital access, skills, self-efficacy, and frustration among older adults; and 3) examine whether digital self-efficacy and frustration may help explain associations between vision impairment and digital access and skills among older adults.

## Methods

### Study Design and Sample

We analyzed a cross-sectional dataset from The National Cancer Institute’s Health Information National Trends Survey (HINTS) 7 [43], which was collected in 2024 (N = 7,278). HINTS is a nationally representative, repeated cross-sectional survey of non-institutionalized U.S. adults aged 18 or older that has been conducted periodically since 2003. HINTS tracks trends in the American public’s need for, access to, and use of health-related information and health-related behaviors, perceptions, and knowledge. The rationale for selecting a cross-sectional design using HINTS 2024 alone is that HINTS 2024 most accurately reflects the latest landscape of digital technology access and use among older adults with vision impairment after the COVID-19 pandemic. Furthermore, the variables of vision impairment, digital self-efficacy, and digital frustration were newly introduced in HINTS 2024, enabling us to examine these constructs together. Data collection for HINTS 2024 occurred from March to September 2024 using paper or web modes. The weighted response rate for HINTS 2024 was 27.3%. Detailed descriptions of HINTS survey design are published elsewhere [43,44]. This study used publicly available, de-identified data from the HINTS website [43]. This study met the criteria for exemption from institutional review board (IRB) review [45]. Given the focus on older adults in this study, we limited respondents aged 60 or older, which follows the World Health Organization’s definition of older adults as “aged 60 or older” [46]. The final sample included 3,149 respondents representing U.S. older adult population aged 60 or older (see the study flow chart in **Multimedia Appendix 1**). We followed the STROBE (Strengthening the Reporting of Observational Studies in Epidemiology) reporting guidelines [47] (**Multimedia Appendix 2**).

### Conceptual Framework and Operational Definitions

We used the conceptual framework (**Figure 1**) guided by the Senior Technology Acceptance Model (STAM) [27] to understand factors predicting digital technology access and use among older adults with vision impairment. STAM extends the Technology Acceptance Model [48] by incorporating age-related health and ability constructs (e.g., self-reported health conditions), as key predictors of technology usage behaviors among older adults. STAM has been widely used in digital tool usability research among older adults, including smartphones [49], wearable monitoring devices [50], and telehealth technology [51]. In this conceptual framework, vision impairment corresponds to the ‘self-reported health conditions,’ digital self-efficacy corresponds to the ‘gerontechnology self-efficacy,’ digital frustration corresponds to the ‘gerontechnology anxiety,’ and digital access and skills correspond to the ‘usage behavior’ in STAM [27]. While digital frustration shares conceptual overlap with gerontechnology anxiety in that both focus on negative emotional responses toward technology use [27,30], the two constructs differ in their temporal focus: digital frustration captures the emotional response during technology use following a failure, whereas gerontechnology anxiety involves anticipatory apprehension before use. Given the absence of a gerontechnology anxiety measure in HINTS 2024, digital frustration was used as the closest available proxy for this construct.

**Figure 1.**
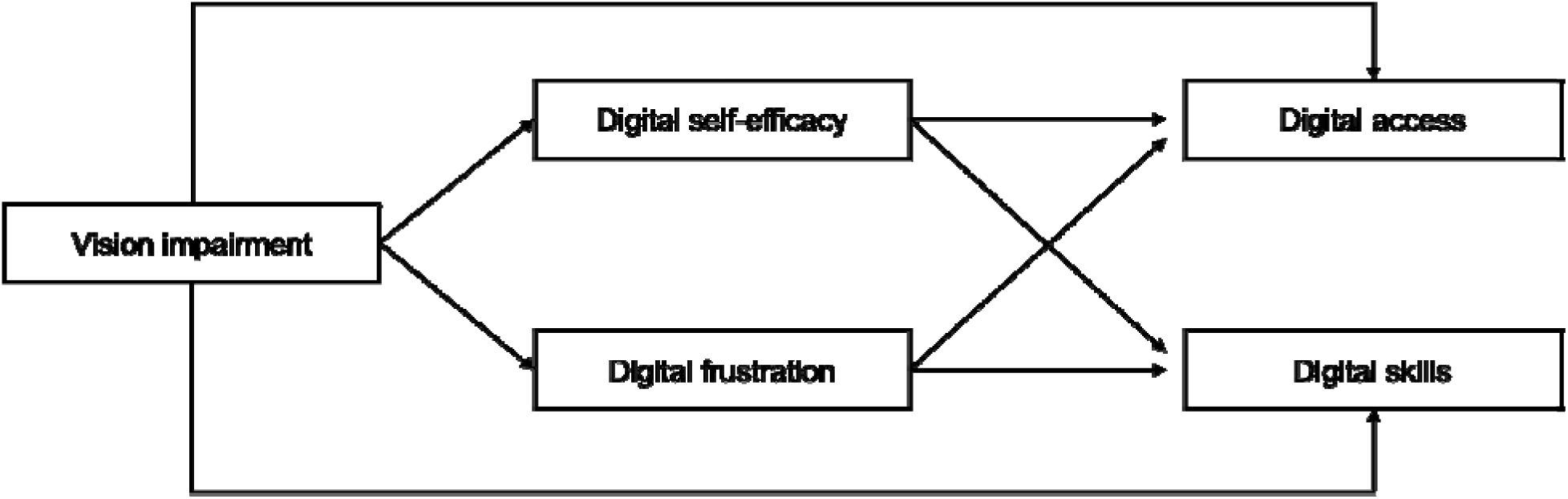
Conceptual framework guided by the Senior Technology Acceptance Model (STAM) among older adults with vision impairment using HINTS 2024 data. **Footnote.** This conceptual framework was guided by Chen and Chan (2014) Senior Technology Acceptance Model (STAM). Vision impairment corresponds to the ‘self-reported health conditions,’ digital self-efficacy corresponds to the ‘gerontechnology self-efficacy,’ digital frustration corresponds to the ‘gerontechnology anxiety,’ and digital access and skills correspond to the ‘usage behavior’ in STAM. Digital frustration is conceptually related to but distinct from gerontechnology anxiety in STAM. While gerontechnology anxiety involves anticipatory apprehension before technology use, digital frustration captures the emotional response during technology use following a failure.

Operational definitions used in this study are as follows. Older adults were defined as people aged 60 or older [46]. Vision impairment refers to a significant limitation of visual capability that cannot be corrected by refractive correction, medication, or surgery [52]. Digital self-efficacy refers to an individual’s perception of their capability to perform tasks related to the use of digital technology [28,30]. Digital frustration refers to an emotional response to a failure associated with using digital technology that impedes goal attainment [33,34]. Digital access refers to the frequency of internet use and the use of digital devices. Digital skills refer to the ability to perform internet-based health-related tasks.

### Measures

#### Vision Impairment

Self-reported visual difficulty was used as a proxy for vision impairment because HINTS does not include objectively measured visual acuity (e.g., Snellen eye chart, logMAR). To assess the presence of vision impairment, participants were asked the following question: “Do any of the following significantly limit your daily activities: visual impairments or blindness?” There were two response options: 1) Yes and 2) No.

#### Digital Access

We assessed five variables of digital access. In terms of the frequency of using the internet, the following question was used: “About how often do you use the Internet, either on a computer, laptop, smartphone or any other device?” There were six response options: 1) More than once per day, 2) About once per day, 3) A few times a week, 4) Less than once per week, 5) Rarely, and 6) Never. We used four questions to ask the use of digital devices as follows: In the last 12 months, did you use: 1) a smartphone; 2) a desktop computer or laptop; 3) a tablet; or 4) a smartwatch or other electronic wearable device (for example and Apple Watch or Fitbit)? There were two response options: 1) Yes and 2) No.

#### Digital Skills

We assessed five variables of digital skills. We used four questions to ask internet-based health-related tasks: In the past 12 months, have you used the Internet to: 1) look for health or medical information; 2) send a message to a healthcare provider or health care providers office; 3) view medical test results; or 4) make an appointment with a healthcare provider? There were two response options: 1) Yes and 2) No. In terms of using telehealth, the following question was used: “In the past 12 months, did you receive care from a doctor or healthcare professional using telehealth?” There were four response options: 1) Yes, by video, 2) Yes, by phone call (voice only with no video), 3) Yes, some by video and some by phone call, and 4) No telehealth visits in the past 12 months.

#### Digital Self-efficacy

We assessed digital self-efficacy with the following question: “How much do you agree or disagree: I can use applications/programs (like Zoom) on my cell phone or computer without asking someone for help.” There were four response options: 1) Strongly agree, 2) Somewhat agree, 3) Somewhat disagree, and 4) Strongly disagree.

#### Digital Frustration

We assessed digital frustration with the following question: “How much do you agree or disagree: I find learning how to use new technology frustrating.” There were four response options: 1) Strongly agree, 2) Somewhat agree, 3) Somewhat disagree, and 4) Strongly disagree.

#### Covariates

We assessed age in years; sex assigned at birth (female, male); race/ethnicity (White, Black or African American, Hispanic, Asian, American Indian or Alaska Native, Native Hawaiian or other Pacific Islander, multiple races); education (5 options: from “1 = Less than High School” to “5 = Post-Baccalaureate Degree”); annual household income (6 options: from “1 = Less than $20,000” to “6 = $100,000 or greater”); and the number of comorbidities (range from 0 to 6: cancer, diabetes, hypertension, heart condition, chronic lung disease, depression or anxiety disorder).

### Statistical Analysis

To ensure results are representative of the U.S. adult population, all analyses were performed using a full-sample weight. Each adult who completed a questionnaire in HINTS 2024 (Cycle 7) received a full-sample weight and a set of 50 replicate weights. The 50 replicate weights were used to calculate accurate standard errors using the ‘delete one’ jackknife (JK1) replication method, in accordance with the HINTS 2024 Methodology Report [43].

Descriptive statistics were used to describe the study population characteristics and prevalence of digital access, skills, self-efficacy, and frustration by the presence of vision impairment. We presented the difference in survey-weighted percentage (95% confidence interval [CI]) for categorical variables and survey-weighted mean for continuous variables.

Survey-weighted multivariable logistic regression models were used to 1) explore factors associated with digital access, skills, self-efficacy, and frustration among older adults with vision impairment and 2) examine the associations between vision impairment and digital access, skills, self-efficacy, and frustration among older adults. All models were adjusted for age, sex, race/ethnicity, education, and the number of comorbidities. These covariates were selected based on a literature review to identify potential confounders [21,24–26]. To facilitate interpretation, we presented adjusted predicted marginal proportions and risk differences between older adults with and without vision impairment. We did not include both education and income as confounders in the same models because they represent related indicators of socioeconomic status. In sensitivity analyses, income was substituted for education to assess the consistency of results for the association between vision impairment and digital access, skills, self-efficacy, and frustration.

After confirming associations in multivariable logistic regression models (*P*<.05), we performed survey-weighted generalized structural equation modeling to examine the potential indirect associations between vision impairment and digital access and skills measures via digital self-efficacy and frustration among older adults. This approach allowed for the simultaneous estimation of two logistic regression models predicting (1) digital self-efficacy and frustration and (2) digital access and skills. The indirect effect of vision impairment on digital access and skills via digital self-efficacy and frustration was evaluated using the Delta method implemented through the ‘nlcom’ command in Stata. This method provides a computationally stable alternative to bootstrapping when using complex survey weights [53]. We reported the indirect effect on the log-odds coefficient with 95% CIs as an estimate of the indirect association in the specified model. All models were adjusted for age, sex, race/ethnicity, education, and the number of comorbidities.

Since the survey-weighted proportions of missing data for most variables were less than 10% (range: 0.0–8.4%), we conducted a complete case analysis. Missing values for the four internet-based digital skills outcomes (i.e., used the internet to look for health information, send a message to a health care provider, view medical test results, and make an appointment with a health care provider) among older adults with vision impairment (range: 18.7–19.1%) were likely attributable to the HINTS 2024 skip pattern that classifies respondents who reported ‘Never’ as ‘Inapplicable,’ rather than solely to item nonresponse. Responses of ‘Missing data (Not Ascertained),’ ‘Missing data (Web partial – Question Never Seen),’ ‘Multiple responses selected in error,’ ‘Question answered in error (Commission Error),’ and ‘Inapplicable’ were treated as missing data and excluded from the analyses. Statistical significance was set at a two-tailed *P* value <.05. Data were analyzed from July 2025 to June 2026 using Stata v19.5 [54].

## Results

### Sample Characteristics

Among a total of 3,149 older adults aged 60 or older (mean [SD] age, 70.7 [10.0] years; 1,773 female [45.6%]; 1,355 male [54.4%]), participants self-identified their race and ethnicity as follows: 404 Hispanic (8.5%), 452 Non-Hispanic Black or African American (11.1%), 1,904 Non-Hispanic White (73.0%), and 192 Non-Hispanic Other (7.4%), which included American Indian, Alaska Native, Asian, multiple races, Native Hawaiian, or other Pacific Islander (**Table 1**). The 1,295 older adults (26.3%) reported having a bachelor’s degree or higher. A total of 223 of 3,149 participants (7.1%) self-reported vision impairment. Older adults with vision impairment were more likely to identify as male (difference, 12.5%; 95% CI, 0.4 to 24.5), earn less income (< $20,000: difference, 14.2%; 95% CI, 3.5 to 24.9), and have more comorbidities (≥4 comorbidities: difference, 12.4%; 95% CI, 3.1 to 21.8) compared with those without visual impairment.

**Table 1.** Sample characteristics among older adults by the presence of vision impairment, Health Information National Trends Survey 2024 (N = 3,149)^a^.

|  | Total older adults<br>(N = 3,149) | Older adults without VI<br>(n = 2,926 [92.9%]) | Older adults with VI<br>(n = 223 [7.1%]) | Difference in weighted means or proportions<br>(95% CI)<sup>b</sup> |
| --- | --- | --- | --- | --- |
| Age, mean (SD), year | 70.7 (10.0) | 70.6 (9.8) | 72.1 (12.8) | 1.5 (-1.1 to 4.1) |
| Missing data, No. | 0 | 0 | 0 |  |
| Sex (at birth), No. (%) |  |  |  |  |
| Female | 1,773 (45.6) | 1,659 (46.3) | 114 (33.9) | -12.5 (-24.5 to -0.4) |
| Male | 1,355 (54.4) | 1,249 (53.7) | 106 (66.1) | 12.5 (0.4 to 24.5) |
| *Missing data, No.* | 21 | 18 | 3 |  |
| Race/ethnicity, No. (%) |  |  |  |  |
| Hispanic | 404 (8.5) | 366 (8.2) | 38 (13.2) | 5.0 (-1.6 to 11.6) |
| Non-Hispanic Black or African American | 452 (11.1) | 422 (11.0) | 30 (11.7) | 0.7 (-7.2 to 8.6) |
| Non-Hispanic White | 1,904 (73.0) | 1,785 (73.9) | 119 (59.6) | -14.2 (-27.9 to -0.6) |
| Non-Hispanic Other<sup>c</sup> | 192 (7.4) | 175 (6.9) | 17 (15.4) | 8.5 (-4.9 to 22.0) |
| *Missing data, No.* | 197 | 178 | 19 |  |
| Education, No. (%) |  |  |  |  |
| ≤ High school graduate | 813 (30.1) | 736 (29.3) | 77 (42.5) | 13.1 (-1.1 to 27.3) |
| Some college | 1,025 (43.6) | 963 (44.1) | 62 (34.9) | -9.2 (-22.8 to 4.5) |
| ≥ Bachelor's degree | 1,295 (26.3) | 1,213 (26.6) | 82 (22.6) | -4.0 (-11.7 to 3.8) |
| *Missing data, No.* | 16 | 14 | 2 |  |
| Annual household income, \$, No. (%) | | | | |
| < 20,000 | 550 (13.9) | 475 (13.1) | 75 (27.3) | 14.2 (3.5 to 24.9) |
| 20,000 to < 50,000 | 873 (28.6) | 800 (28.4) | 73 (32.0) | 3.6 (-7.9 to 15.2) |
| 50,000 to < 75,000 | 489 (18.2) | 459 (18.5) | 30 (14.1) | -4.3 (-12.2 to 3.5) |
| > 75,000 | 1,001 (39.3) | 971 (40.0) | 30 (26.6) | -13.5 (-27.7 to 0.7) |
| *Missing data, No.* | 236 | 221 | 15 |  |
| Number of comorbidities, No. (%) |  |  |  |  |
| 0 | 535 (19.8) | 514 (20.6) | 21 (6.7) | -13.9 (-18.9 to -9.0) |
| 1 | 909 (28.2) | 863 (28.5) | 46 (22.8) | -5.7 (-18.0 to 6.7) |
| 2 | 879 (27.4) | 813 (27.5) | 66 (26.6) | -0.9 (-14.1 to 12.3) |
| 3 | 510 (17.6) | 466 (17.2) | 44 (25.2) | 8.0 (-5.5 to 21.6) |
| ≥4 | 242 (7.0) | 201 (6.2) | 41 (18.7) | 12.4 (3.1 to 21.8) |
| *Missing data, No.* | 74 | 69 | 5 |  |

Abbreviation: CI: confidence interval, SD: standard deviation, VI: vision impairment
<sup>a</sup> Numbers are unweighted; % and mean represents survey-weighted estimates.
<sup>b</sup> Difference in survey-weighted means for continuous variables and survey-weighted proportions for categorical variables.
<sup>c</sup> Other includes participants who self-identify as American Indian, Alaska Native, Asian, multiple races, Native Hawaiian, or other Pacific Islander.

### Survey-weighted Prevalences of Digital Access, Skills, Self-efficacy, and Frustration

**Figure 2** and **Table 2** presented the weighted prevalence of digital access, skills, self-efficacy, and frustration among older adults by the presence of vision impairment in HINTS 2024 sample. The weighted prevalence of digital access, skills, self-efficacy, and frustration among older adults with vision impairment were as follows: used the internet daily (65.6%; 95% CI, 53.5% to 75.9%); used a smartphone (79.5%; 95% CI, 66.8% to 88.2%), computer (58.9%; 95% CI, 46.4% to 70.3%), tablet (32.8%; 95% CI, 20.7% to 47.7%), and wearable device (16.5%; 95% CI, 8.4% to 29.8%) in the past 12 months; used the internet to look for health information (76.8%; 95% CI, 63.1% to 86.5%), message to a health care provider (56.6%; 95% CI, 43.3% to 69.0%), view medical test results (68.0%; 95% CI, 56.2% to 77.9%), make an appointment with a health care provider (44.0%; 95% CI, 31.0% to 57.8%) in the past 12 months; used any telehealth in the past 12 months (32.8%; 95% CI, 21.1% to 47.1%); had digital self-efficacy (45.2%; 95% CI, 33.8% to 57.2%); and had digital frustration (82.1%; 95% CI, 68.9% to 90.4%).

**Figure 2.**
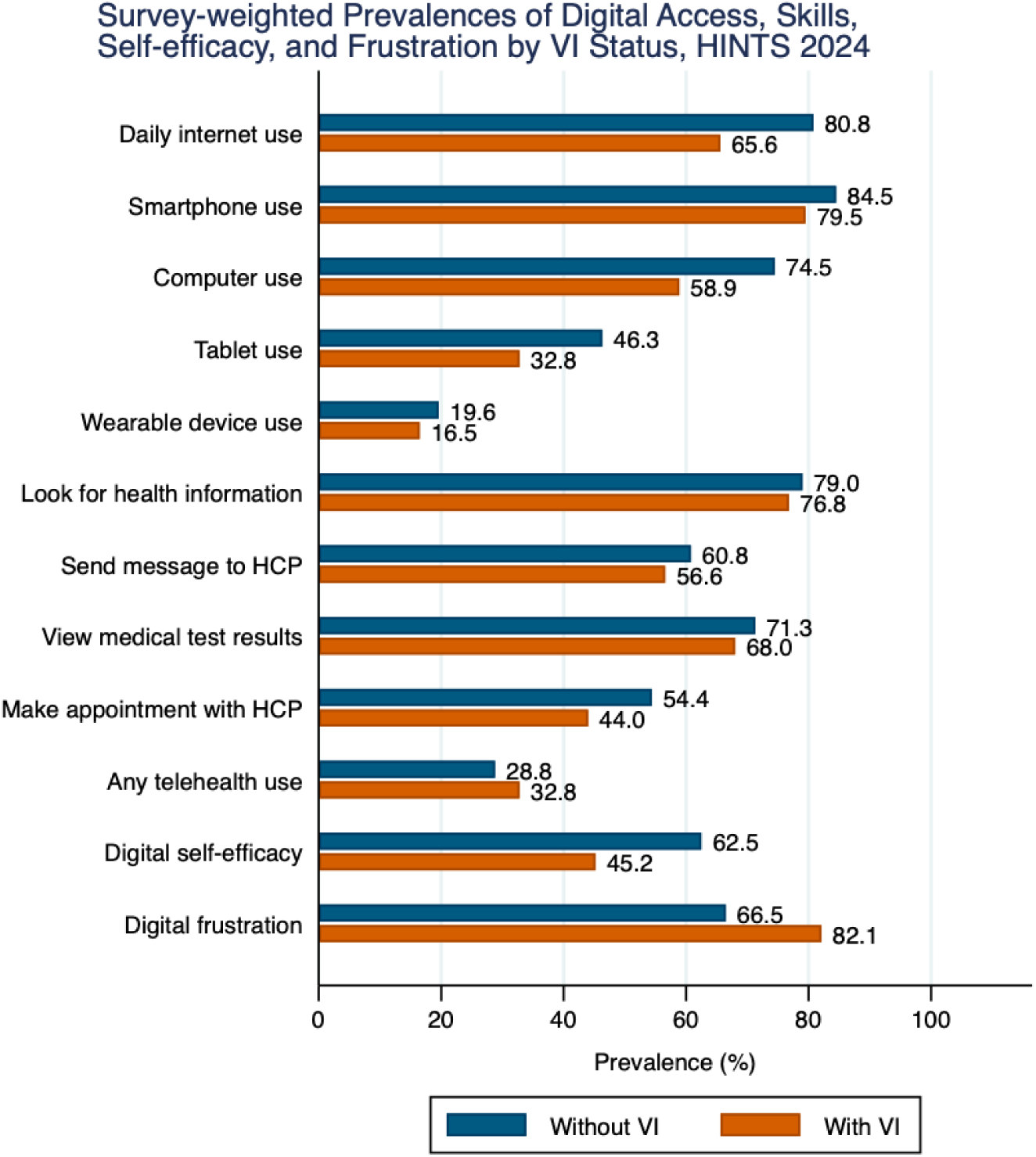
Survey-weighted prevalences of digital access, skills, self-efficacy, and frustration among older adults by vision impairment status, Health Information National Trends Survey 2024 (N = 3,149)^a,b^. Abbreviation: HCP: health care provider, HINTS: Health Information National Trends Survey, VI: vision impairment

**Table 2.**
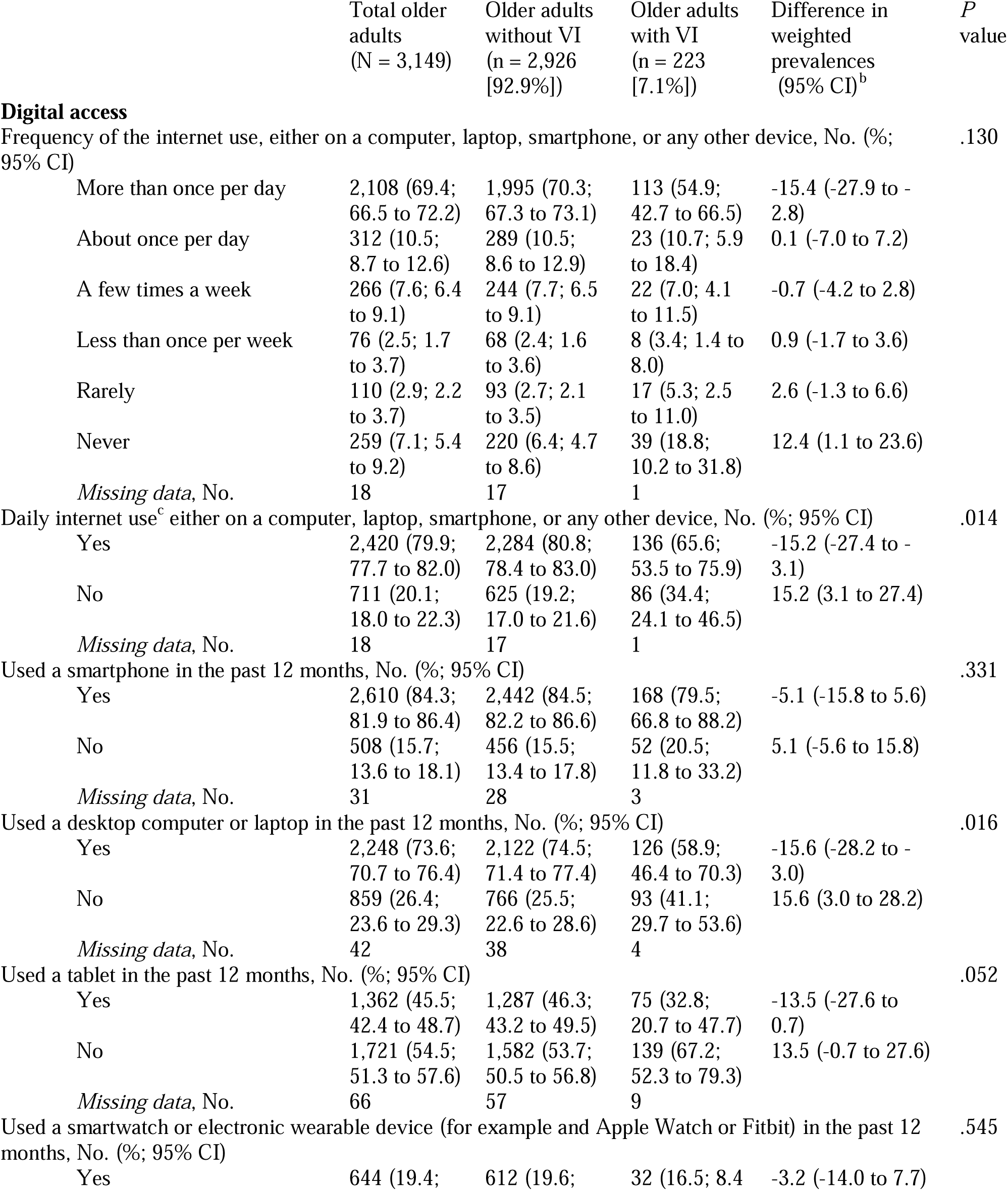
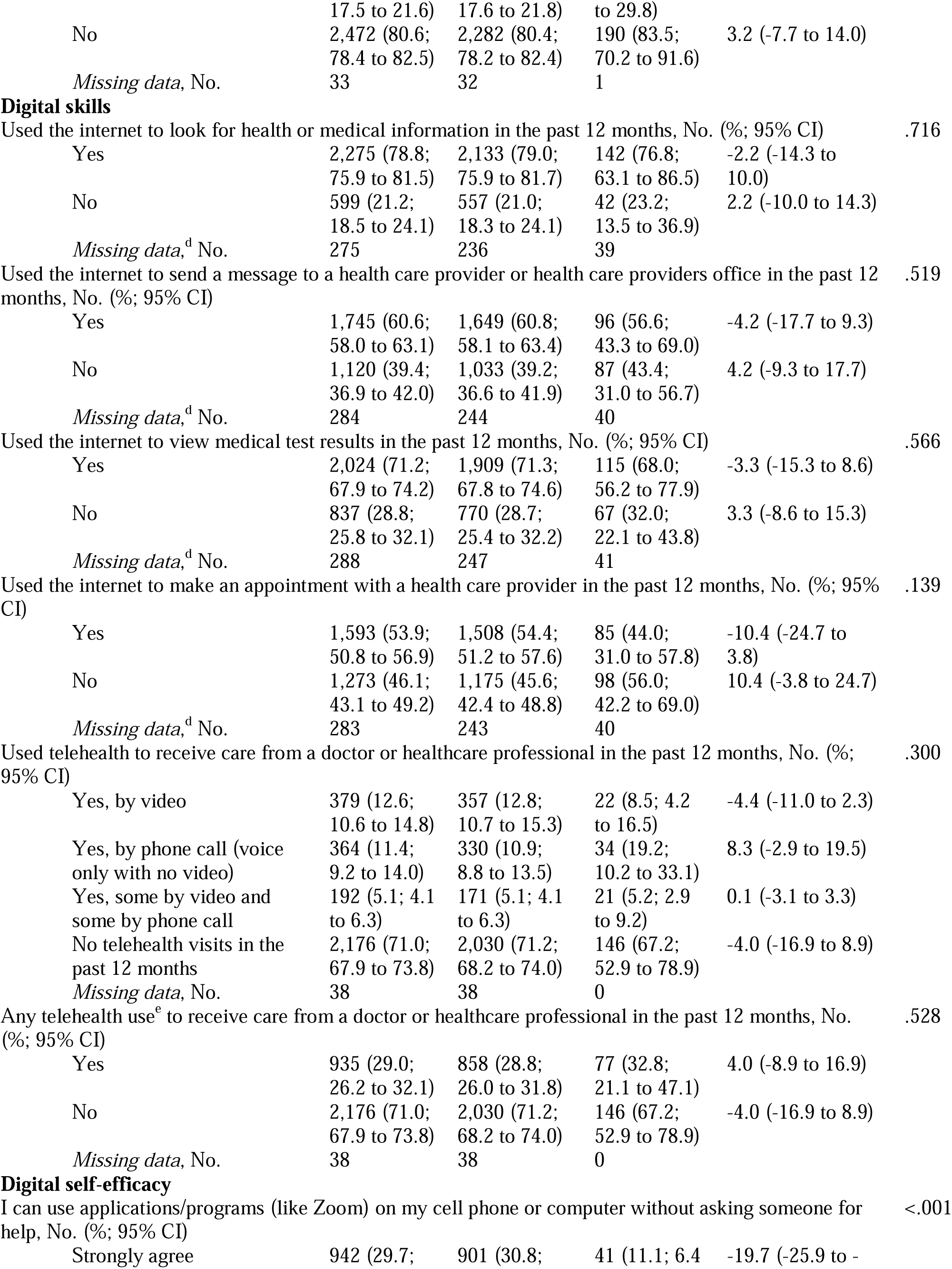
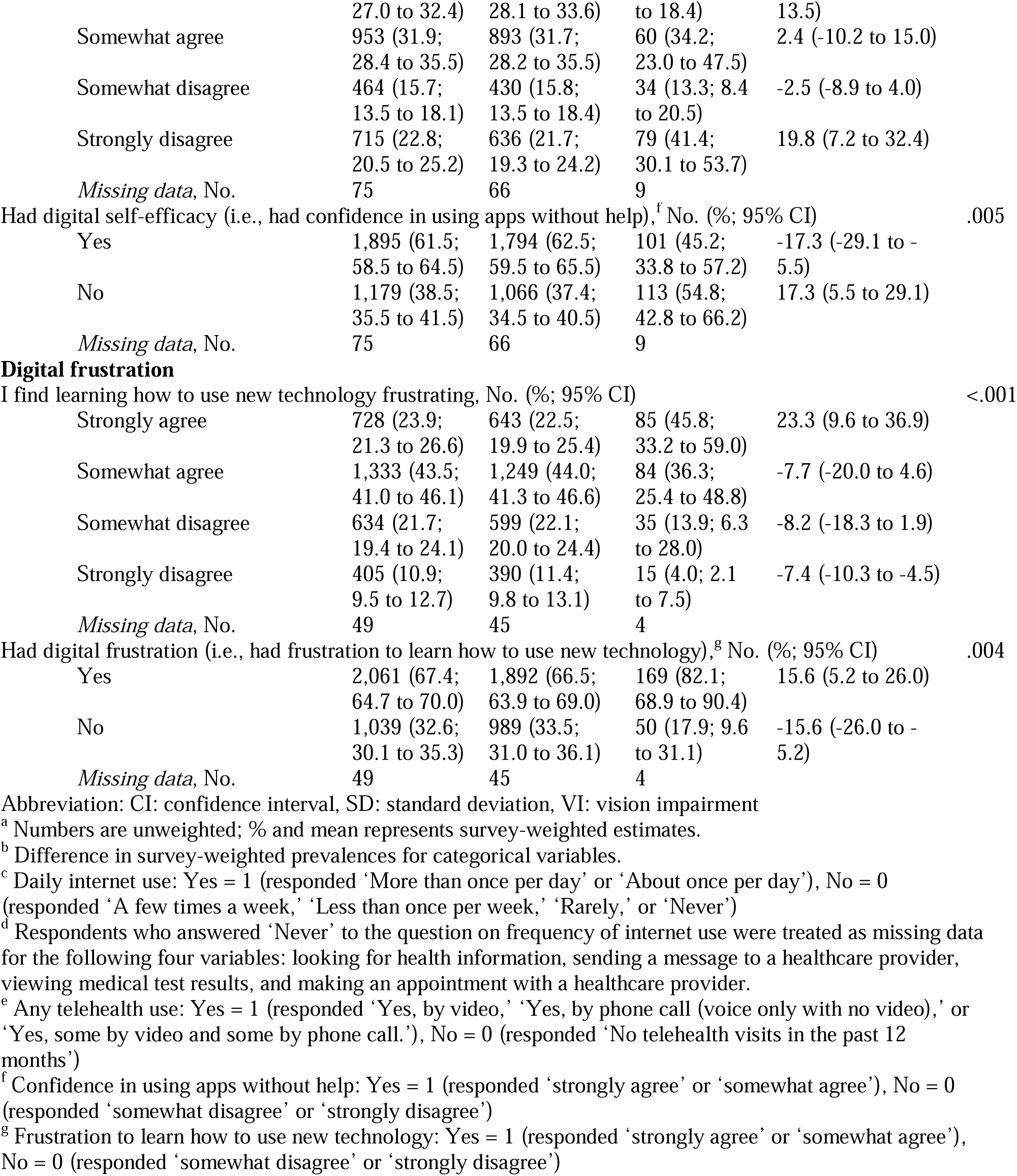
Survey-weighted prevalences of digital access, skills, self-efficacy, and frustration among older adults by the presence of vision impairment, Health Information National Trends Survey 2024 (N = 3,149)^a,b^.

Compared with those without vision impairment, older adults with vision impairment had lower prevalence of using the internet daily (-15.2%: 95% CI, -27.4% to -3.1%) and using a computer in the past 12 months (-15.6%: 95% CI, -28.2% to -3.0%). Furthermore, differences existed between older adults without and with vision impairment in digital self-efficacy (-17.3%; 95% CI, -29.1% to -5.5%) and digital frustration (15.6%; 95% CI, 5.2% to 26.0%). There were no differences in the other digital access and skills variables by the presence of vision impairment.

### Multivariable Logistic Regression Analyses

In survey-weighted multivariable logistic regression analyses among older adults with vision impairment (**Table 3** and **Table 4**), older age was associated with lower odds of daily internet use (odds ratio [OR], 0.84; 95% CI, 0.79 to 0.90), smartphone use (OR, 0.85; 95% CI, 0.75 to 0.97), wearable device use (OR, 0.88; 95% CI, 0.79 to 0.97), and using the internet to send a message to a healthcare provider (OR, 0.87; 95% CI, 0.80 to 0.93). Older adults who self-identified as racial and ethnic minority groups (i.e., American Indian, Alaska Native, Asian, Black/African American, Hispanic, multiple races, Native Hawaiian, or other Pacific Islander) had lower odds of daily internet use (OR, 0.15; 95% CI, 0.05 to 0.50) and using the internet to send a message to a healthcare provider (OR, 0.17; 95% CI, 0.04 to 0.73) compared with Non-Hispanic White older adults. No other associations were noted between factors and digital access, skills, self-efficacy, and frustration variables.

**Table 3.**
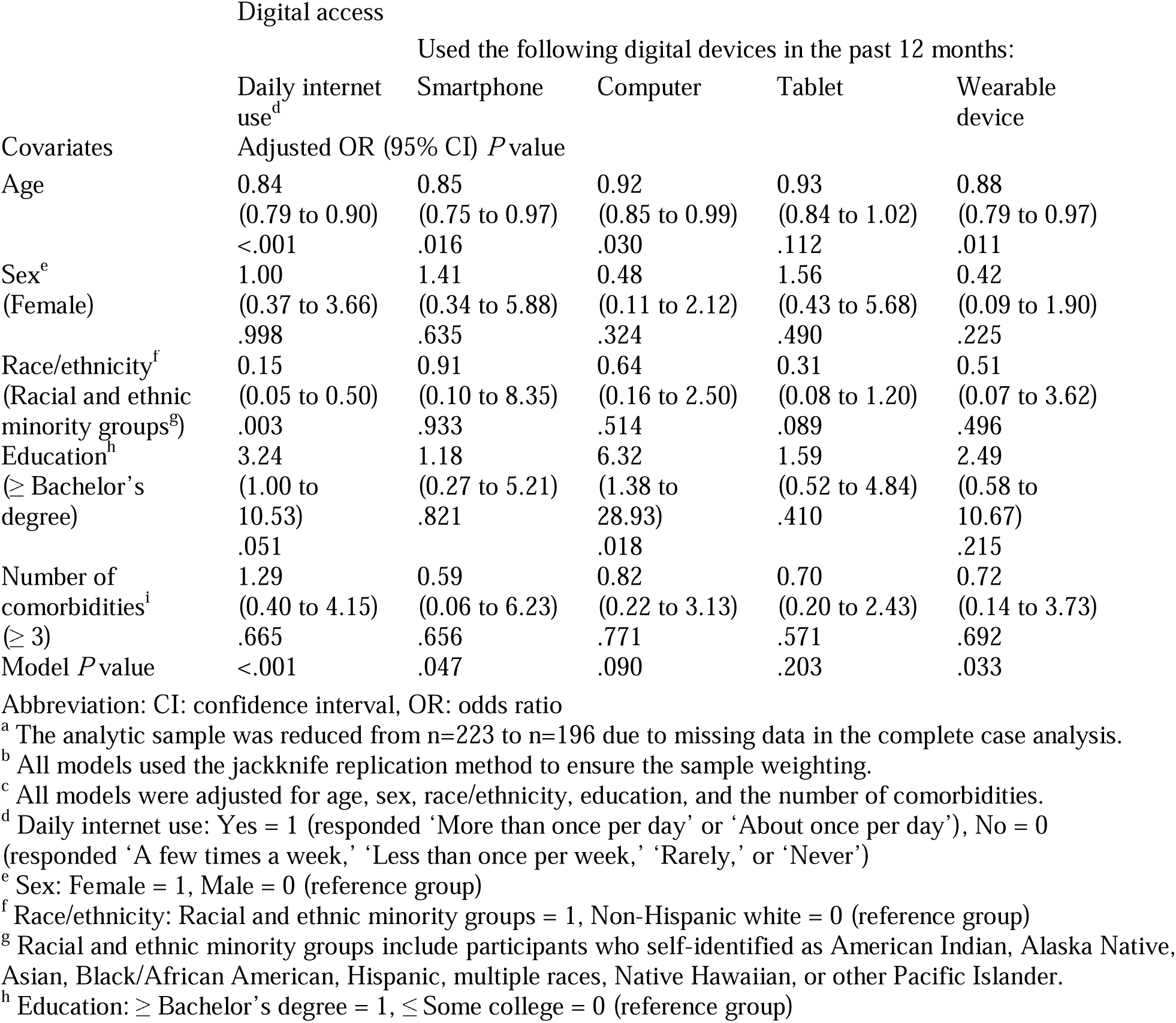
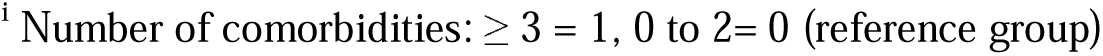
Survey-weighted multivariable logistic regression analyses on digital access among older adults with vision impairment (n = 196)^a-c^.

| Covariates | Digital access |  |  |  |  |
| --- | --- | --- | --- | --- | --- |
|  | Used the following digital devices in the past 12 months: |  |  |  |  |
|  | Daily internet use<sup>d</sup> | Smartphone | Computer | Tablet | Wearable device |
|  | Adjusted OR (95% CI) | P value |  |  |  |
| Age | 0.84<br>(0.79 to 0.90)<br><.001 | 0.85<br>(0.75 to 0.97)<br>.016 | 0.92<br>(0.85 to 0.99)<br>.030 | 0.93<br>(0.84 to 1.02)<br>.112 | 0.88<br>(0.79 to 0.97)<br>.011 |
| Sex<sup>e</sup><br>(Female) | 1.00<br>(0.37 to 3.66)<br>.998 | 1.41<br>(0.34 to 5.88)<br>.635 | 0.48<br>(0.11 to 2.12)<br>.324 | 1.56<br>(0.43 to 5.68)<br>.490 | 0.42<br>(0.09 to 1.90)<br>.225 |
| Race/ethnicity<sup>f</sup><br>(Racial and ethnic minority groups<sup>g</sup>) | 0.15<br>(0.05 to 0.50)<br>.003 | 0.91<br>(0.10 to 8.35)<br>.933 | 0.64<br>(0.16 to 2.50)<br>.514 | 0.31<br>(0.08 to 1.20)<br>.089 | 0.51<br>(0.07 to 3.62)<br>.496 |
| Education<sup>h</sup><br>(≥ Bachelor's degree) | 3.24<br>(1.00 to 10.53)<br>.051 | 1.18<br>(0.27 to 5.21)<br>.821 | 6.32<br>(1.38 to 28.93)<br>.018 | 1.59<br>(0.52 to 4.84)<br>.410 | 2.49<br>(0.58 to 10.67)<br>.215 |
| Number of comorbidities<sup>i</sup><br>(≥ 3) | 1.29<br>(0.40 to 4.15)<br>.665 | 0.59<br>(0.06 to 6.23)<br>.656 | 0.82<br>(0.22 to 3.13)<br>.771 | 0.70<br>(0.20 to 2.43)<br>.571 | 0.72<br>(0.14 to 3.73)<br>.692 |
| Model P value | <.001 | .047 | .090 | .203 | .033 |

Abbreviation: CI: confidence interval, OR: odds ratio
<sup>a</sup> The analytic sample was reduced from n=223 to n=196 due to missing data in the complete case analysis.
<sup>b</sup> All models used the jackknife replication method to ensure the sample weighting.
<sup>c</sup> All models were adjusted for age, sex, race/ethnicity, education, and the number of comorbidities.
<sup>d</sup> Daily internet use: Yes = 1 (responded 'More than once per day' or 'About once per day'), No = 0 (responded 'A few times a week,' 'Less than once per week,' 'Rarely,' or 'Never')
<sup>e</sup> Sex: Female = 1, Male = 0 (reference group)
<sup>f</sup> Race/ethnicity: Racial and ethnic minority groups = 1, Non-Hispanic white = 0 (reference group)
<sup>g</sup> Racial and ethnic minority groups include participants who self-identified as American Indian, Alaska Native, Asian, Black/African American, Hispanic, multiple races, Native Hawaiian, or other Pacific Islander.
<sup>h</sup> Education: ≥ Bachelor's degree = 1, ≤ Some college = 0 (reference group)
<sup>i</sup> Number of comorbidities: $\geq 3 = 1$ , 0 to 2 = 0 (reference group)

**Table 4.**
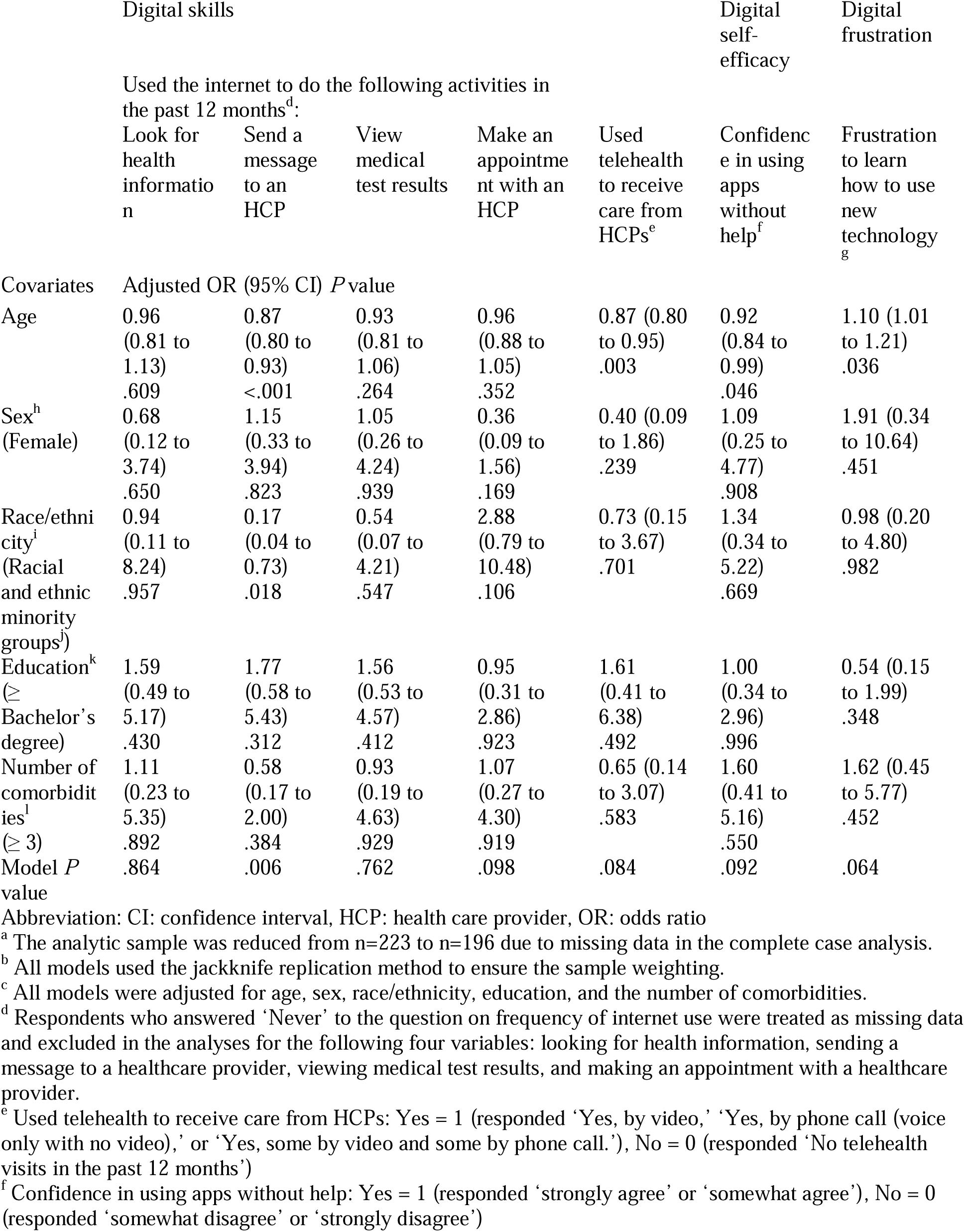
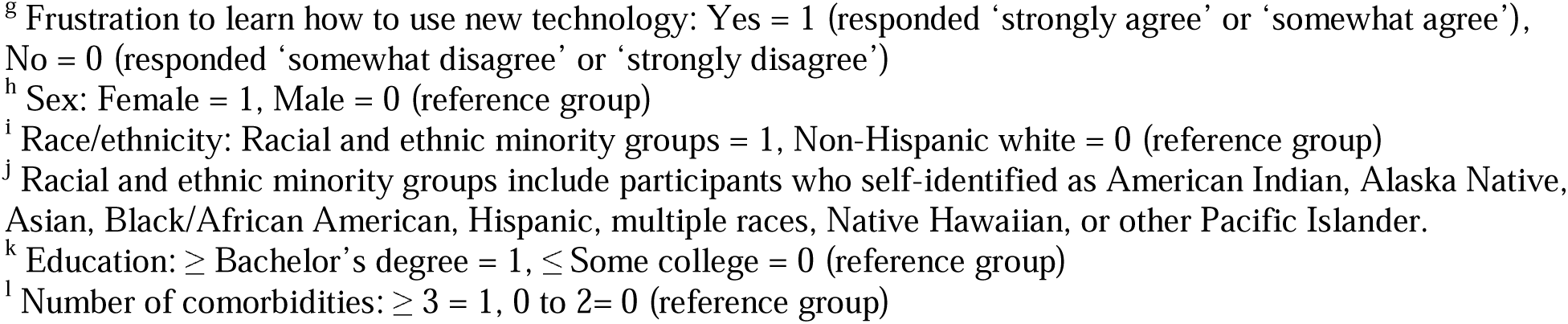
Survey-weighted multivariable logistic regression analyses on digital skills, self-efficacy, and frustration among older adults with vision impairment (n = 196)^a-c^.

|  | Digital skills |  |  |  |  | Digital self-efficacy | Digital frustration |
| --- | --- | --- | --- | --- | --- | --- | --- |
|  | Used the internet to do the following activities in the past 12 months<sup>d</sup>: |  |  |  |  |  |  |
|  | Look for health information | Send a message to an HCP | View medical test results | Make an appointment with an HCP | Used telehealth to receive care from HCPs<sup>e</sup> | Confidence in using apps without help<sup>f</sup> | Frustration to learn how to use new technology<sup>g</sup> |
| Covariates | Adjusted OR (95% CI) *P* value |  |  |  |  |  |  |
| Age | 0.96<br>(0.81 to 1.13)<br>.609 | 0.87<br>(0.80 to 0.93)<br><.001 | 0.93<br>(0.81 to 1.06)<br>.264 | 0.96<br>(0.88 to 1.05)<br>.352 | 0.87 (0.80 to 0.95)<br>.003 | 0.92<br>(0.84 to 0.99)<br>.046 | 1.10 (1.01 to 1.21)<br>.036 |
| Sex<sup>h</sup><br>(Female) | 0.68<br>(0.12 to 3.74)<br>.650 | 1.15<br>(0.33 to 3.94)<br>.823 | 1.05<br>(0.26 to 4.24)<br>.939 | 0.36<br>(0.09 to 1.56)<br>.169 | 0.40 (0.09 to 1.86)<br>.239 | 1.09<br>(0.25 to 4.77)<br>.908 | 1.91 (0.34 to 10.64)<br>.451 |
| Race/ethnicity<sup>i</sup><br>(Racial and ethnic minority groups<sup>j</sup>) | 0.94<br>(0.11 to 8.24)<br>.957 | 0.17<br>(0.04 to 0.73)<br>.018 | 0.54<br>(0.07 to 4.21)<br>.547 | 2.88<br>(0.79 to 10.48)<br>.106 | 0.73 (0.15 to 3.67)<br>.701 | 1.34<br>(0.34 to 5.22)<br>.669 | 0.98 (0.20 to 4.80)<br>.982 |
| Education<sup>k</sup><br>(≥ Bachelor's degree) | 1.59<br>(0.49 to 5.17)<br>.430 | 1.77<br>(0.58 to 5.43)<br>.312 | 1.56<br>(0.53 to 4.57)<br>.412 | 0.95<br>(0.31 to 2.86)<br>.923 | 1.61<br>(0.41 to 6.38)<br>.492 | 1.00<br>(0.34 to 2.96)<br>.996 | 0.54 (0.15 to 1.99)<br>.348 |
| Number of comorbidities<sup>l</sup><br>(≥ 3) | 1.11<br>(0.23 to 5.35)<br>.892 | 0.58<br>(0.17 to 2.00)<br>.384 | 0.93<br>(0.19 to 4.63)<br>.929 | 1.07<br>(0.27 to 4.30)<br>.919 | 0.65 (0.14 to 3.07)<br>.583 | 1.60<br>(0.41 to 5.16)<br>.550 | 1.62 (0.45 to 5.77)<br>.452 |
| Model *P* value | .864 | .006 | .762 | .098 | .084 | .092 | .064 |

Abbreviation: CI: confidence interval, HCP: health care provider, OR: odds ratio
<sup>a</sup> The analytic sample was reduced from n=223 to n=196 due to missing data in the complete case analysis.
<sup>b</sup> All models used the jackknife replication method to ensure the sample weighting.
<sup>c</sup> All models were adjusted for age, sex, race/ethnicity, education, and the number of comorbidities.
<sup>d</sup> Respondents who answered 'Never' to the question on frequency of internet use were treated as missing data and excluded in the analyses for the following four variables: looking for health information, sending a message to a healthcare provider, viewing medical test results, and making an appointment with a healthcare provider.
<sup>e</sup> Used telehealth to receive care from HCPs: Yes = 1 (responded 'Yes, by video,' 'Yes, by phone call (voice only with no video),' or 'Yes, some by video and some by phone call.'), No = 0 (responded 'No telehealth visits in the past 12 months')
<sup>f</sup> Confidence in using apps without help: Yes = 1 (responded 'strongly agree' or 'somewhat agree'), No = 0 (responded 'somewhat disagree' or 'strongly disagree')
<sup>g</sup> Frustration to learn how to use new technology: Yes = 1 (responded ‘strongly agree’ or ‘somewhat agree’), No = 0 (responded ‘somewhat disagree’ or ‘strongly disagree’)
<sup>h</sup> Sex: Female = 1, Male = 0 (reference group)
<sup>i</sup> Race/ethnicity: Racial and ethnic minority groups = 1, Non-Hispanic white = 0 (reference group)
<sup>j</sup> Racial and ethnic minority groups include participants who self-identified as American Indian, Alaska Native, Asian, Black/African American, Hispanic, multiple races, Native Hawaiian, or other Pacific Islander.
<sup>k</sup> Education: ≥ Bachelor’s degree = 1, ≤ Some college = 0 (reference group)
<sup>l</sup> Number of comorbidities: $\geq 3 = 1$ , 0 to 2 = 0 (reference group)

In survey-weighted multivariable logistic regression models examining the associations between vision impairment and digital access, skills, self-efficacy, and frustration (**Table 5**), older adults with vision impairment had lower odds of daily internet use (OR, 0.60; 95% CI, 0.37 to 0.99) and digital self-efficacy (i.e., confidence in using apps without help) (OR, 0.53; 95% CI, 0.32 to 0.86) compared with those without vision impairment. No other associations were noted between vision impairment and digital access, skills, or frustration variables. In sensitivity analyses adjusting for income instead of education, results were consistent with the primary models other than no association between vision impairment with daily internet use (**Multimedia Appendix 3**).

**Table 5.**
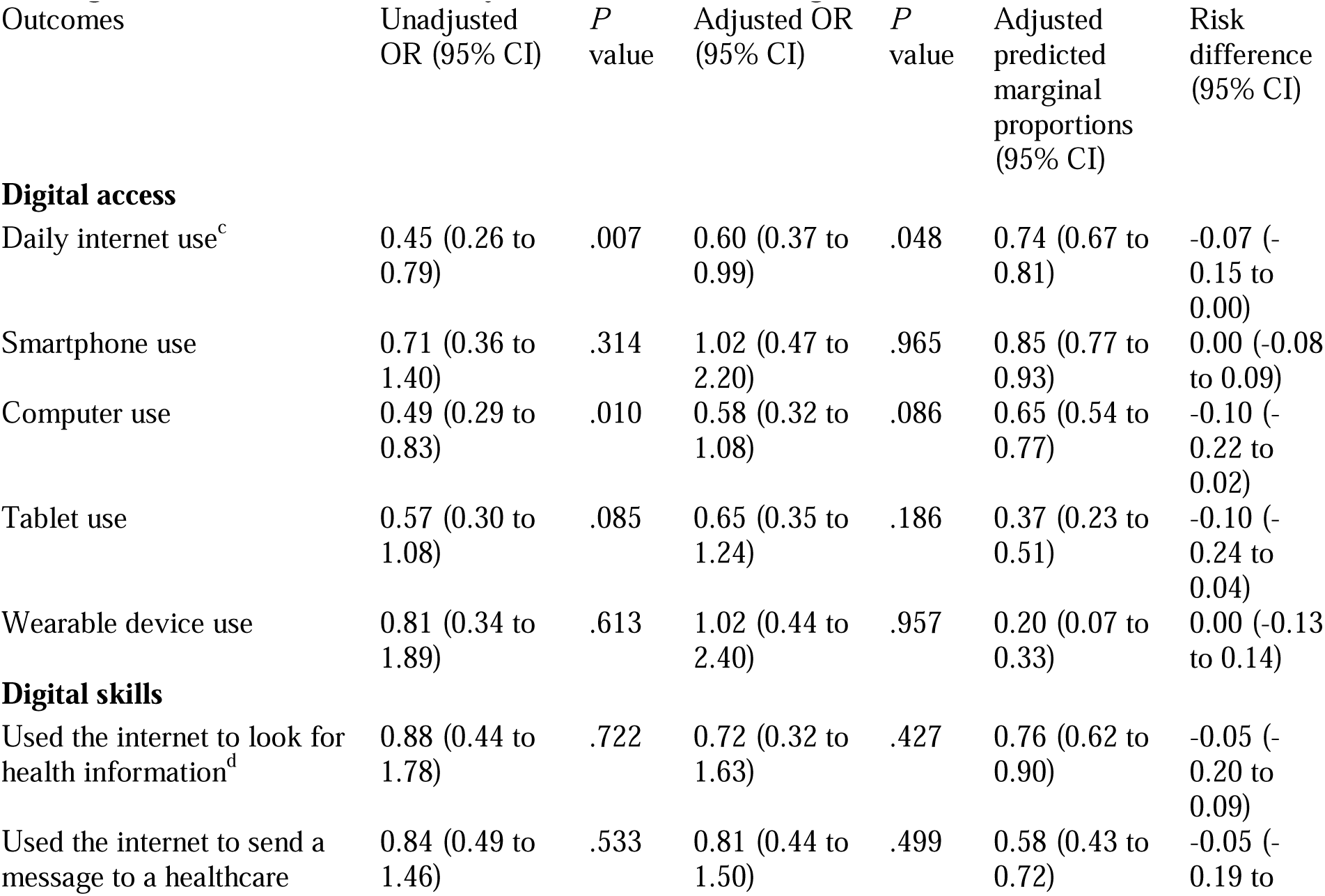
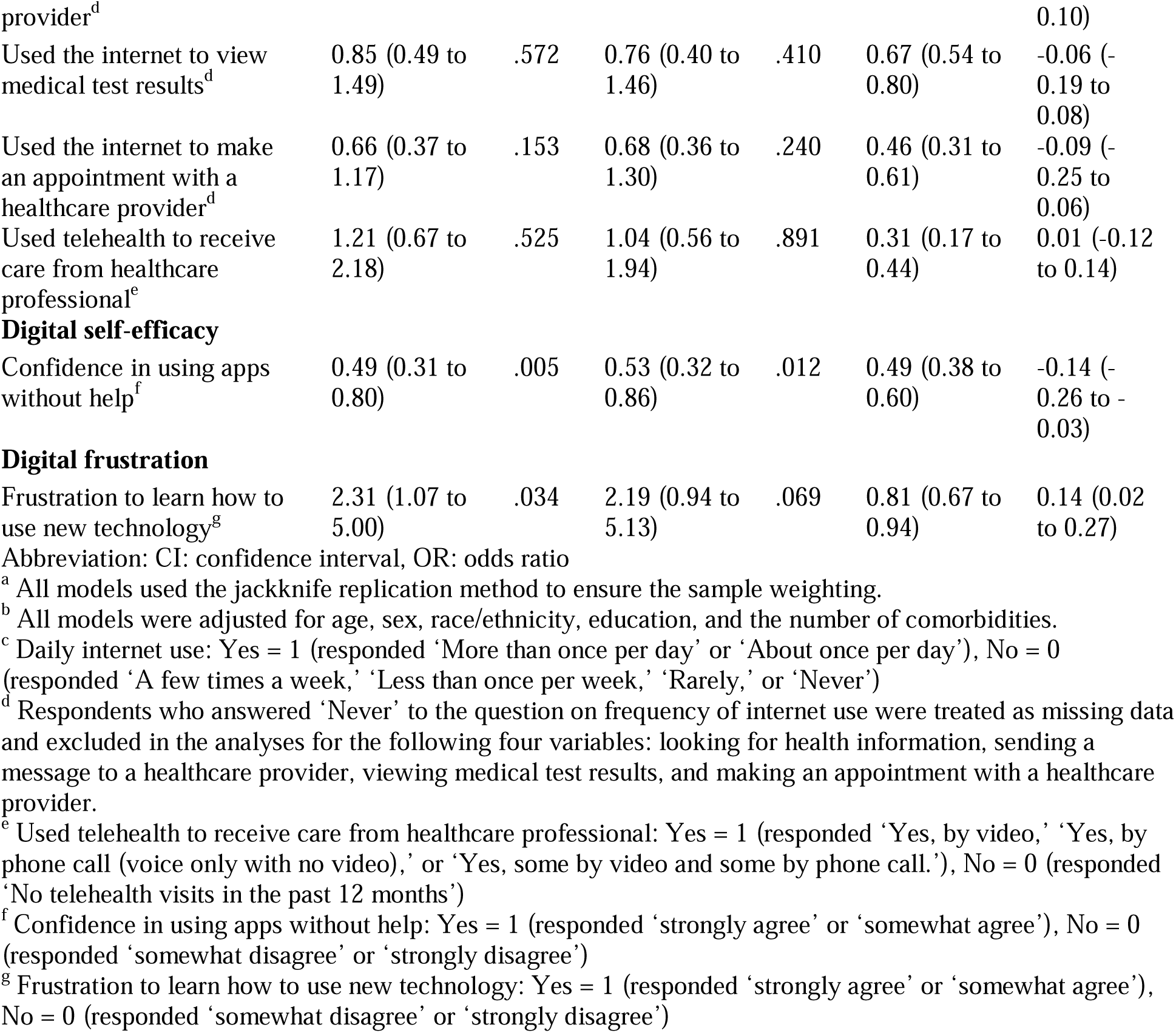
Survey-weighted multivariable logistic regression: Association between vision impairment and digital access, skills, self-efficacy, and frustration among older adults (n = 2,848)^a,b^.

Since vision impairment was associated with both daily internet use and digital self-efficacy (i.e., confidence in using apps without help; **Table 5**), we examined a potential indirect association between vision impairment and daily internet use via digital self-efficacy among older adults. In survey-weighted multivariable logistic regression models (**Table 6**), digital self-efficacy was associated with higher odds of daily internet use (OR, 2.95; 95% CI, 2.04 to 4.26). Generalized structural equation modeling showed an indirect association between vision impairment and daily internet use via digital self-efficacy (coefficient, -0.68; 95% CI, -1.24 to -0.12).

**Table 6.** Survey-weighted multivariable logistic regression: indirect association between vision impairment and daily internet use via digital self-efficacy among older adults (n = 2,848)^a-c^.

| Path | Unadjusted OR<br>(95% CI) | P value | Adjusted OR<br>(95% CI) | P value |
| --- | --- | --- | --- | --- |
| VI – Digital self-efficacy<sup>d</sup> | 0.49 (0.31 to 0.80) | .005 | 0.53 (0.32 to 0.86) | .012 |
| Digital self-efficacy<sup>d</sup> – Daily internet use<sup>e</sup> | 3.48 (2.49 to 4.86) | <.001 | 2.95 (2.04 to 4.26) | <.001 |
| VI – Daily internet use<sup>e</sup> | 0.45 (0.26 to 0.79) | .007 | 0.60 (0.37 to 0.99) | .048 |

**Abbreviations.** CI: confidence interval, OR: odds ratio, VI: vision impairment.
<sup>a</sup> All models used the jackknife replication method to ensure the sample weighting.
<sup>b</sup> All models were adjusted for age, sex, race/ethnicity, education, and the number of comorbidities.
<sup>c</sup> All multivariable logistic regression models in Table 6 achieved statistical significance (Model P value, $P < .05$ )
<sup>d</sup> Confidence in using apps without help: Yes = 1 (responded ‘strongly agree’ or ‘somewhat agree’), No = 0 (responded ‘somewhat disagree’ or ‘strongly disagree’)
<sup>e</sup> Daily internet use: Yes = 1 (responded ‘More than once per day’ or ‘About once per day’), No = 0 (responded ‘A few times a week,’ ‘Less than once per week,’ ‘Rarely,’ or ‘Never’)

## Discussion

### Principal Findings

This cross-sectional study using HINTS 2024 examined the associations between vision impairment and digital access, skills, self-efficacy, and frustration among older adults in the U.S. To the best of our knowledge, this is the first study to show that 1) older age and racial and ethnic minority groups were associated with lower odds of daily internet use and using the internet to send a message to a healthcare provider among older adults with vision impairment, 2) vision impairment was associated with lower odds of digital self-efficacy among older adults, 3) digital self-efficacy was associated with higher odds of daily internet use among older adults, and 4) there was evidence of an indirect association between vision impairment and daily internet use via digital self-efficacy among older adults. The findings suggest that reduced digital self-efficacy may be an important factor in understanding the observed association between vision impairment and daily internet use among older adults. However, these findings should be interpreted with caution, given the cross-sectional design and the small sample of older adults with vision impairment (n=223).

The prevalence of digital access among older adults with vision impairment in this study was consistent with the findings from a cross-sectional study of the National Health and Aging Trends Study (NHATS) 2021 by Thomas et al. in 2024 [24]. Both studies found lower prevalences of computer and tablet access and use among older adults with vision impairment compared with those without vision impairment (computer use: 58.9% vs. 74.5% in this study; computer ownership and knowledge: 61.5% vs. 78.0% in Thomas et al.; tablet use: 32.8% vs. 46.3% in this study; tablet ownership and knowledge: 40.1% vs. 56.0% in Thomas et al.). However, direct comparison should be interpreted with caution, as NHATS 2021 assessed ownership and knowledge of these digital devices, whereas HINTS 2024 assessed actual use in the past 12 months. Interestingly, telehealth use showed a different pattern between the two studies. In our study, older adults with vision impairment had slightly higher telehealth use than those without vision impairment (32.8% vs. 28.8%), whereas Thomas et al. found lower telehealth use among those with vision impairment (42.6% vs. 46.2%). However, neither study found statistically significant differences in telehealth use by the presence of vision impairment. The overall lower telehealth use observed in our study compared to Thomas et al. [24] may reflect the decline in telehealth use following the acute phase of the COVID-19 pandemic, as NHATS data were collected in 2021 during a period of high telehealth adoption. These findings suggest that disparities in digital access still persist among older adults with vision impairment in the U.S., underscoring the need for targeted interventions to improve digital access in this population.

Older age was associated with lower odds of using the internet daily, a smartphone, a wearable device, and using the internet to send a message to a healthcare provider among older adults with vision impairment. These findings are consistent with a repeated cross-sectional study using HINTS 2017–2020 data among older adults in the U.S. [21]. Several mechanisms may explain why older age was associated with lower digital access and skills among older adults with vision impairment. In terms of physical health, older age is generally associated with greater cognitive decline [55], including reduced working memory, slower information processing speed, and decreased attention, which may be associated with difficulties in learning how to use new digital technology. In addition, older age is associated with a greater number of comorbidities [56] and worsening self-rated health [57], which may be associated with fatigue attributed to chronic diseases, contributing to limited energy for digital technology use. Furthermore, consistent with the STAM framework [27], some data suggest that older age is associated with negative attitudes toward digital technology [58] and greater technology anxiety [59], which may inhibit the intention and actual usage behaviors of digital technology among older adults. At the same time, in the current sample about 80% of older adults without vision impairment and over 60% of those with vision impairment reported using the internet on a daily basis, suggesting the necessity to further evaluate age as well as other related factors that may affect digital engagement in this population.

Racial and ethnic minority groups, including Black race and Hispanic ethnicity, were also associated with lower odds of daily internet use and using the internet to send a message to a healthcare provider among older adults with vision impairment. These findings underscore the existence of disproportionate racial and ethnic disparities in digital access and skills among older adults with vision impairment. A prior study demonstrated similar findings, showing that Black and Hispanic American older adults were less likely to access the internet and have a smartphone or tablet compared with White American older adults [21]. Older adults who self-identified as racial and ethnic minority groups may be exposed to economic hardship and structural barriers [60], which hinder their ability to purchase internet broadband and digital devices to access digital health care. According to the “Rhizomatic Digital Ecosystem Framework [61],” Affordability—defined as acknowledging that many individuals and families may be unable to pay for internet service, devices, and digital resources even when they are technically available and adequate for their needs— is noted as a key factor to understand digital exclusion. Thus, ensuring affordability is critical for enabling equitable access to digital health care in this population.

Vision impairment was associated with lower odds of digital self-efficacy, digital self-efficacy was associated with higher odds of daily internet use, and there was evidence of an indirect association between vision impairment and daily internet use via digital self-efficacy among older adults. Our findings indicate that vision impairment may further exacerbate the digital access divide among older adults. These findings highlight the importance of further investigating enhancing digital self-efficacy as a modifiable factor to increase digital engagement among older adults with vision impairment.

To understand potential explanations for the observed associations, three factors may contribute to low digital self-efficacy among older adults with vision impairment. First, inaccessible interfaces, including small fonts, low color contrasts, and complex navigation, may lead to repeated operational failures [10], which may subsequently reduce digital self-efficacy. According to Bandura’s social cognitive theory [28], repeated failures in performing a task significantly reduce one’s self-efficacy, defined as one’s belief in their capabilities to perform a specific task, increasing perceived powerlessness and behavioral avoidance of the task. This process is consistent with the learned helplessness theory proposed by Seligman [62], which suggests that repeated uncontrollable failures lead individuals to develop an expectation of future failure, reducing motivation to attempt the task in the future. One qualitative study empirically supports these two theories, showing that older adults perceive fear and shame of making mistakes with digital technology, which was described as one barrier to the use of digital technology among older adults [63]. These theories collectively underpin the notion that older adults with vision impairment may experience lower digital self-efficacy due to repeated technical errors attributed to inaccessible digital interfaces.

Second, the digital environment may impose significant cognitive overload among older adults with vision impairment. Cognitive overload refers to a state in which cognitive demands exceed one’s available cognitive resources [64–66]. Cognitive overload can lead to severe mental fatigue [67–69]. Older adults with vision impairment often experience high cognitive demands due to the necessity to compensate for reduced visual input [67,70]. They may experience cognitive overload because of the need to rely on other senses to substitute for visual input, use of assistive devices, and unique memorization techniques to retain information [67]. One cross-sectional study [69] showed that concentration difficulty is associated with greater fatigue among adults with vision impairment, suggesting that people with vision impairment may experience significant cognitive overload and fatigue due to limited visual inputs. In the context of digital technology use, using a digital device daily can be a challenge for older adults with vision impairment because they need to allocate substantial working memory to address cognitive demands. For example, using the screen reader on a laptop computer requires simultaneous processing of auditory information and execution of commands, imposing substantial demands on working memory. Furthermore, such dual-task processing may be particularly challenging for older adults who experience age-related cognitive decline. The significant cognitive overload may make even minor technical difficulties feel insurmountable hurdles, subsequently contributing to ‘technostress,’ defined as stress created by an inability to effectively adapt to new information and communication technology [71,72]. One empirical study [73] showed that individuals under high technostress experience greater cognitive overload, slower performance speed, and more errors than those under low technostress. Thus, cognitive overload may further reduce digital self-efficacy among older adults with vision impairment, creating barriers to digital engagement in this population.

Third, the cumulative impact of these factors may reduce a sense of autonomy among older adults with vision impairment in an increasingly digital society. Smartphone proficiency has become a proxy for independence and social participation. When family and friends communicate exclusively via smartphone applications, older adults with vision impairment who cannot navigate these digital platforms without assistance may feel a sense of social exclusion. The necessity to seek consistent help to perform basic digital tasks could diminish one’s sense of autonomy and increase a feeling of dependency on others. The process could potentially contribute to ‘internalized ableism,’ defined as that a person with a disability (or disabilities) internalizes negative stereotypes, prejudice, and discrimination against disabilities [74,75], and ‘internalized ageism,’ defined as that an older adult internalizes negative stereotypes, prejudice, and discrimination against old age, aging process, and older adults [76–78]. Internalized ableism and ageism may contribute to older adults with vision impairment perceiving themselves as burdensome or incapable and feeling shame in seeking help to use digital technology. One focus group study [79] suggested that internalized ageism may lead to avoidance of using digital technology among older adults with low confidence in its use. Furthermore, one cross-sectional study using the Health and Retirement Study (HRS) 2016 data [80] reported that negative self-perception of aging was associated with lower prevalence of internet use among older adults. Prior meta-analysis showed that internalized ableism was associated with reduced self-efficacy among adults with disabilities [81]. Internalized ageism was also associated with lower self-esteem among older adults [82]. These findings suggest that internalized ableism and ageism may reduce digital self-efficacy, which may reduce motivation to engage with digital technology among older adults with vision impairment. Therefore, the digital divide may not only represent a gap in access to digital technologies, but may also affect one’s sense of identity, where older adults with vision impairment may perceive a sense of inferiority due to their perceived inability to participate in digital platforms. In summary, these potential factors, including repeated technical errors, cognitive overload, technostress, and internalized ableism and ageism, may contribute to reduced digital self-efficacy and subsequent digital disengagement among older adults with vision impairment. Future studies are needed to further examine whether repeated technical errors, cognitive overload, technostress, and internalized ableism and ageism are associated with reduced digital self-efficacy and digital disengagement in this population.

Vision impairment was not associated with digital frustration among older adults. This finding may reflect limited statistical power to detect the association between vision impairment and digital frustration given the small sample size of older adults with vision impairment (n=223), as suggested by the wide confidence interval (OR, 2.19; 95% CI, 0.94 to 5.13). Despite the limited precision, the magnitude of this point estimate suggests a potential impact, indicating that older adults with vision impairment may be at greater risk of experiencing digital frustration compared with those without vision impairment. According to Bandura’s social cognitive theory [28], individuals with low self-efficacy may respond differently depending on their outcome expectancies and others’ performance in using digital technology. When individuals hold low self-efficacy beliefs and low outcome expectancies and see others like them fail to use digital technology, they are less likely to initiate or sustain effort toward using digital technology, as they perceive little worth in doing so, ultimately leading to digital disengagement. In contrast, when individuals hold low self-efficacy beliefs and high outcome expectancies and see others like them successfully use digital technology, they may experience frustration and self-criticism as they recognize the value of the task but doubt their ability to perform it. This cognitive and behavioral process is also theoretically aligned with STAM [27], in which self-reported health conditions (i.e., vision impairment) are associated with technology anxiety, contributing to reduced digital technology use. It is plausible that poor accessibility in digital interfaces may generate repeated operational failures and cognitive overload, which may be associated with digital frustration among older adults with vision impairment. Together, these theoretical perspectives suggest the importance of assessing digital frustration to better understand low digital engagement in this population. Future research with an adequate sample size is needed to examine this association.

### Implications for Practice

These preliminary findings provide useful knowledge to guide accessible interface designs and personalized coaching programs to enhance digital self-efficacy and digital technology use among older adults with vision impairment. Accessible interface designs may reduce repeated operational failures and cognitive overload in this population. For example, simplifying user interfaces and pre-configuring devices that the most important apps already set up with accessibility features may help remove the initial barriers to using digital devices. Personalized coaching programs that fit the user’s needs and pace to address their specific challenges with a certain app or digital device (e.g., Zoom’s keyboard shortcuts, screen magnification, VoiceOver/TalkBack) may help improve digital self-efficacy. According to Bandura’s social cognitive theory [28], there are four sources of self-efficacy: 1) enactive mastery experience (i.e., prior success in performing a task strengthens the motivation to try a task next time); 2) vicarious experience (i.e., observation, learning, and comparison to others similar to oneself succeed increases one’s confidence); 3) verbal persuasion (i.e., receiving realistic encouragement from others helps overcome self-doubt); and 4) physiologic feedback (i.e., managing stress and emotional responses when facing challenges). Approaches to these sources of self-efficacy may be useful to design novel digital training programs for improving digital self-efficacy. Regarding ‘enactive mastery experience,’ module-based learning to break down complex tasks into manageable small modules (e.g., in the case of using the Zoom app, ‘joining a Zoom meeting’, ‘turning on your camera and microphone’) may help older adults with vision impairment experience continued small success accomplishments and promote confidence in using apps and digital devices. One empirical study [83] is consistent with this approach, showing that mastery of past experience (i.e., prior successful experience in performing a specific task) was associated with higher self-efficacy among older adults. Regarding ‘vicarious experience,’ peer support groups for older adults, where they can learn from someone who understands their experiences and can share their frustrations and successes may reduce feelings of isolation and build their confidence. Regarding ‘verbal persuasion,’ positive feedback from family, friends, and health care providers may facilitate continued digital engagement among older adults with vision impairment. One cross-sectional study [84] found that social support mediated the relationship between digital health literacy and self-efficacy among older adults, suggesting that verbal persuasion through social support may be useful to enhance self-efficacy for seeking and using electronic health information. Regarding ‘physiologic feedback,’ mindfulness-based techniques or structured relaxation exercises before using digital technology may help manage technostress and digital frustration in this population. Furthermore, initiatives to increase awareness that learning new technology is a challenge for everyone, regardless of age or ability, may reduce internalized ageism and ableism. In summary, accessible interface designs, personalized coaching programs, and initiatives on sources of self-efficacy may be beneficial to enhance digital self-efficacy and digital engagement in this population.

### Strengths and Limitations

There are several strengths in our study. First, we used a nationally representative sample of U.S. older adults, which enhanced the external validity of the findings. Second, rich data on types of digital devices and internet-based health-related tasks provided us with a granular understanding of digital access and use among older adults with vision impairment. Third, digital self-efficacy and frustration measures gave us a better understanding of the indirect determinants of digital technology use behaviors in this population.

However, several limitations should be acknowledged. First, given the cross-sectional design, we were unable to establish temporality between vision impairment and digital access, skills, self-efficacy, and frustration. The results of the indirect association analysis should be interpreted with caution, as the findings may be hypothesis-generating rather than confirmatory. Furthermore, in sensitivity analysis substituting household income for education, there was no association between vision impairment and daily internet use. This finding suggests that the observed association may be sensitive to the socioeconomic variables included in the model, and should therefore be interpreted cautiously. Future studies are needed to better disentangle the potentially distinct roles of educational attainment, financial resources, and digital self-efficacy in shaping internet use among older adults with vision impairment. Second, a low response rate (27.3%) for HINTS 2024 may have introduced selection bias. Third, the self-reported measure of vision impairment may have introduced misclassification of vision impairment. Future research is needed to incorporate objective measures of visual acuity and underlying eye conditions. In addition, self-reported measures of digital access and skills may have introduced recall bias. Future research is needed to objectively assess actual digital technology usage patterns, including frequency and duration, in this population. Fourth, since the digital self-efficacy and frustration measures each include only one item, it is unclear whether these single items fully capture the conceptual breadth of these constructs, and we were unable to assess their reliability and validity. Furthermore, the digital self-efficacy measure in HINTS 2024 only focused on device-specific (i.e., cell phone or computer) and software-specific (e.g., Zoom) domains, which may have failed to capture other key constructs of digital self-efficacy, including content creation, addressing safety concerns, and problem-solving behaviors [30]. Fifth, since the HINTS 2024 dataset did not include cognitive function measures, it is unclear whether cognitive function is associated with digital technology use in this population. Sixth, missing data on internet-based health-related skills may have introduced selection bias to overestimate the prevalence of digital skills. Seventh, the HINTS 2024 dataset did not include measures of digital device ownership. Finally, we were unable to assess other key constructs in STAM [27], including perceived usefulness, perceived easy of use, and the attitude and intention toward digital technology use. Future research is needed to explore these constructs in STAM.

## Conclusions

This cross-sectional study using HINTS 2024 examined the associations between vision impairment and digital access, skills, self-efficacy, and frustration among U.S. older adults. Older age and racial and ethnic minority groups were associated with lower odds of daily internet use and using the internet to send a message to a healthcare provider among older adults with vision impairment. Vision impairment was associated with lower daily internet use and lower digital self-efficacy among older adults. Digital self-efficacy was associated with higher daily internet use. There was evidence of an indirect association between vision impairment and daily internet use via digital self-efficacy among older adults. The findings suggest that reduced digital self-efficacy may be an important factor in understanding the observed association between vision impairment and daily internet use among older adults. Interventions to improve digital self-efficacy, including accessible interface designs and personalized coaching programs, may help enhance digital engagement and bridge the digital divide among older adults with vision impairment. Future research is needed to clarify the relationships between digital self-efficacy and repeated operational failures, cognitive overload, technostress, and internalized ableism and ageism in this population.

## Supporting information

Multimedia Appendix

## Data Availability

All data produced are available online at the Health Information National Trends Survey (HINTS) website: https://hints.cancer.gov/

## Acknowledgments

None.

## Data Availability

The datasets analyzed during this study are publicly available from the Health Information National Trends Survey (HINTS) website (https://hints.cancer.gov/).

## Abbreviations

CI: confidence interval
HINTS: Health Information National Trends Survey
OR: odds ratio
SD: standard deviation
STAM: Senior Technology Acceptance Model
VI: vision impairment

## References

1. GBD 2019 Blindness and Vision Impairment Collaborators; Vision Loss Expert Group of the Global Burden of Disease Study. Trends in prevalence of blindness and distance and near vision impairment over 30 years: an analysis for the Global Burden of Disease Study. Lancet Glob Health. 2021;9(2):e130–e143. doi:10.1016/S2214-109X(20)30425-3

2. Blindness and vision impairment. World Health Organization. URL: https://www.who.int/news-room/fact-sheets/detail/blindness-and-visual-impairment [accessed 2026-06-06]

3. Killeen OJ, De Lott LB, Zhou Y, et al. Population Prevalence of Vision Impairment in US Adults 71 Years and Older: The National Health and Aging Trends Study. JAMA Ophthalmol. 2023;141(2):197–204. doi:10.1001/jamaophthalmol.2022.5840

4. Rahmati M, Smith L, Boyer L, et al. Vision impairment and associated daily activity limitation: A systematic review and meta-analysis. PLoS One. 2025;20(1):e0317452. doi:10.1371/journal.pone.0317452

5. Parravano M, Petri D, Maurutto E, et al. Association Between Visual Impairment and Depression in Patients Attending Eye Clinics: A Meta-analysis. JAMA Ophthalmol. 2021;139(7):753–761. doi:10.1001/jamaophthalmol.2021.1557

6. Shang X, Zhu Z, Wang W, et al. The Association between Vision Impairment and Incidence of Dementia and Cognitive Impairment: A Systematic Review and Meta-analysis. Ophthalmology. 2021;128(8):1135–1149. doi:10.1016/j.ophtha.2020.12.029

7. Ehrlich JR, Ramke J, Macleod D, et al. Association between vision impairment and mortality: a systematic review and meta-analysis. Lancet Glob Health. 2021;9(4):e418–e430. doi:10.1016/S2214-109X(20)30549-0

8. Assi L, Chamseddine F, Ibrahim P, et al. A Global Assessment of Eye Health and Quality of Life: A Systematic Review of Systematic Reviews. JAMA Ophthalmol. 2021;139(5):526–541. doi:10.1001/jamaophthalmol.2021.0146

9. Gajarawala SN, Pelkowski JN. Telehealth Benefits and Barriers. J Nurse Pract. 2021;17(2):218–221. doi: 10.1016/j.nurpra.2020.09.013

10. Hamideh Kerdar S, Bächler L, Kirchhoff BM. The accessibility of digital technologies for people with visual impairment and blindness: a scoping review. Discover Computing. 2024;27(1):24. doi:10.1007/s10791-024-09460-7

11. Remillard ET, Koon LM, Mitzner TL, et al. Everyday Challenges for Individuals Aging With Vision Impairment: Technology Implications. Gerontologist. 2024;64(6):gnad169. doi:10.1093/geront/gnad169

12. Hepburn J, Williams L, McCann L. Barriers to and Facilitators of Digital Health Technology Adoption Among Older Adults With Chronic Diseases: Updated Systematic Review. JMIR Aging. 2025;8:e80000. doi:10.2196/80000

13. Wilson J, Heinsch M, Betts D, et al. Barriers and facilitators to the use of e-health by older adults: a scoping review. BMC Public Health. 2021;21(1):1556. doi:10.1186/s12889-021-11623-w

14. Birati Y, Tzemah-Shahar R. Barriers to Digital Health Adoption in Older Adults: Scoping Review Informed by Innovation Resistance Theory. J Med Internet Res. 2026;28:e75591. doi:10.2196/75591

15. van Dijk JA. The Digital divide. Cambridge: Polity Press; 2020.

16. van Dijk J, Hacker K. The digital divide as a complex and dynamic phenomenon. Info Soc 2003;19:315–26. doi:10.1080/01972240309487

17. Scheerder A, Van Deursen A, Van Dijk J. Determinants of Internet skills, uses and outcomes. A systematic review of the second-and third-level digital divide. Telematics and informatics. 2017;34(8):1607–1624. doi:10.1016/j.tele.2017.07.007

18. van Deursen AJ, van Dijk JA. The first-level digital divide shifts from inequalities in physical access to inequalities in material access. New Media Soc. 2019;21(2):354–375. doi:10.1177/1461444818797082

19. Hargittai E. Second-level digital divide: differences in people’s online skills. First Monday. 2002;7(4):10. doi:10.5210/fm.v7i4.942

20. Deursen AJ, van Dijk JA. Measuring internet skills. Int J Hum Comput Interact. 2010;26(10):891–916. doi:10.1080/10447318.2010.496338

21. Yang R, Gao S, Jiang Y. Digital divide as a determinant of health in the U.S. older adults: prevalence, trends, and risk factors. BMC Geriatr. 2024;24(1):1027. doi:10.1186/s12877-024-05612-y

22. Gu J, Qiu Y, Liu J, et al. Prevalence and influencing factors of digital smart devices use among older adults: a systematic review and meta-analysis. Arch Gerontol Geriatr. 2026;140:106034. doi:10.1016/j.archger.2025.106034

23. Yang S, Cha MJ, van Kessel R, et al. Understanding Inequalities in Mobile Health Utilization Across Phases: Systematic Review and Meta-Analysis. J Med Internet Res. 2025;27:e71349. doi:10.2196/71349

24. Thomas J, Almidani L, Swenor BK, et al. Digital Technology Use Among Older Adults With Vision Impairment. JAMA Ophthalmol. 2024;142(5):445–452. doi:10.1001/jamaophthalmol.2024.0467

25. Lange A, Lange N, Jagiełło K, et al. Use of Digital Technology Among Older Adults in Poland With and Those Without Near Visual Impairment: Cross-Sectional Study. J Med Internet Res. 2025;27:e68947. doi:10.2196/68947

26. Takesue A, Hiratsuka Y, Kondo K, et al. Association Between Visual Impairment and Daily Internet Use Among Older Japanese Individuals: Cross-Sectional Questionnaire Study. JMIR Form Res. 2024;8:e58729. doi:10.2196/58729

27. Chen K, Chan AHS. Gerontechnology acceptance by elderly Hong Kong Chinese: A senior technology acceptance model (STAM). Ergonomics. 2024;57:635–652. doi:10.1080/00140139.2014.895855

28. Bandura A. Self-efficacy: The exercise of control. New York: W.H. Freeman, 1997.

29. Chen K, Lou VWQ. Measuring Senior Technology Acceptance: Development of a Brief, 14-Item Scale. Innov Aging. 2020;4(3):igaa016. doi:10.1093/geroni/igaa016

30. Ulfert-Blank AS, Schmidt I. Assessing digital self-efficacy: Review and scale development. Computers & Education. 2022;191:104626 doi:10.1016/j.compedu.2022.104626

31. Zhang H, Zhao Y, Li J, et al. Developing digital health literacy among nursing interns: A sequential mediation model of preceptor support, self-efficacy, and technology acceptance. Nurse Educ Today. 2026. doi:10.1016/j.nedt.2026.107130

32. Chang L, Polansky M, Yi D, et al. Exploring health care learners’ perceptions of AI integration in the curriculum: a survey tool and findings. BMC Med Educ. 2026;26(1):630. doi:10.1186/s12909-026-08931-3

33. Hadlington L, Scase MO. End-user frustrations and failures in digital technology: exploring the role of Fear of Missing Out, Internet addiction and personality. Heliyon. 2018;4(11):e00872. doi:10.1016/j.heliyon.2018.e00872

34. Bessiere K, Newhagen JE, Robinson JP, et al. A model for computer frustration: The role of instrumental and dispositional factors on incident, session, and post-session frustration and mood. Computers in human behavior. 2006;22(6):941–961.doi:10.1016/j.chb.2004.03.015

35. Chen Y, Yuan J, Li C, et al. Innovative adoption model for digital health technologies among elderly with chronic diseases: integrating Unified Theory of Acceptance and Use of Technology and Knowledge-Attitude-Practice model in a survey of 1222 patients in Shanghai. BMJ Open. 2026;16(3):e105529. doi:10.1136/bmjopen-2025-105529

36. Goodarzi F, Barati M, Rahbar S, et al. Factors associated with intention to use rehabilitation assistive technologies in older adults. Disabil Rehabil Assist Technol. 2026;21(2):604–615. doi:10.1080/17483107.2025.2579038

37. Tsai CY, Tzeng YL, Teng YK, et al. The mediating role of technology anxiety in the impact of loneliness on technology behavioral intention among community-dwelling older adults. PLoS One. 2025;20(5):e0321144. doi:10.1371/journal.pone.0321144

38. An J, Wan K, Xiang Z, et al. Effects of privacy concerns on older adults’ discontinuous usage intention: the chain mediating effect of technology anxiety and the moderating role of perceived price value. Front Public Health. 2025;13:1543409. doi:10.3389/fpubh.2025.1543409

39. An J, Zhu X, Wan K, et al. Older adults’ self-perception, technology anxiety, and intention to use digital public services. BMC Public Health. 2024;24(1):3533. doi:10.1186/s12889-024-21088-2

40. Chen Y, Yuan J, Shi L, et al. Understanding the Role of Technology Anxiety in the Adoption of Digital Health Technologies (DHTs) by Older Adults with Chronic Diseases in Shanghai: An Extension of the Unified Theory of Acceptance and Use of Technology (UTAUT) Model. Healthcare (Basel). 2024;12(14):1421. doi:10.3390/healthcare12141421

41. Birati Y, Tzemah-Shahar R. Barriers to Digital Health Adoption in Older Adults: Scoping Review Informed by Innovation Resistance Theory. J Med Internet Res. 2026;28:e75591. doi:10.2196/75591

42. Wang T, Dong Q, Sun Q, et al. Trajectories and Influencing Factors of Online Health Information-Seeking Behaviors Among Community-Dwelling Older Adults: Longitudinal Mixed Methods Study. J Med Internet Res. 2025;27:e77549. doi:10.2196/77549

43. Health Information National Trends Survey (HINTS). National Cancer Institute. URL: https://hints.cancer.gov/ [accessed 2026-06-06]

44. Nelson DE, Kreps GL, Hesse BW, et al. The Health Information National Trends Survey (HINTS): development, design, and dissemination. J Health Commun. 2004;9(5):443–84. doi:10.1080/10810730490504233

45. Institutional Review Board (IRB) Approvals for the Health Information National Trends Survey (HINTS). National Cancer Institute. URL: https://hints.cancer.gov/about-hints/institutional-review-board.aspx [accessed 2026-06-06]

46. Ageing and health. World Health Organization. URL: https://www.who.int/news-room/fact-sheets/detail/ageing-and-health [accessed 2026-06-06]

47. von Elm E, Altman DG, Egger M, et al. The Strengthening the Reporting of Observational Studies in Epidemiology (STROBE) statement: guidelines for reporting observational studies. J Clin Epidemiol. 2008;61(4):344–349. doi:10.1016/j.jclinepi.2007.11.008

48. Davis FD, Bagozzi RP, Warshaw PR. User acceptance of computer technology: A comparison of two theoretical models. Management Science. 1989;35:982–1003. doi:10.1287/mnsc.35.8.982

49. Petrovčič A, Peek S, Dolničar V. Predictors of Seniors’ Interest in Assistive Applications on Smartphones: Evidence from a Population-Based Survey in Slovenia. Int J Environ Res Public Health. 2019;16(9):1623. doi:10.3390/ijerph16091623

50. Li J, Ma Q, Chan AH, et al. Health monitoring through wearable technologies for older adults: Smart wearables acceptance model. Appl Ergon. 2019;75:162–169. doi:10.1016/j.apergo.2018.10.006

51. van Houwelingen CT, Ettema RG, Antonietti MG, Kort HS. Understanding Older People’s Readiness for Receiving Telehealth: Mixed-Method Study. J Med Internet Res. 2018;20(4):e123. doi:10.2196/jmir.8407

52. MedGen-visual impairment. National Library of Medicine. URL: https://www.ncbi.nlm.nih.gov/medgen/777085 [accessed 2026-06-06]

53. Falk DS, Vazquez CE, Handique S. Psychological Distress Mediates the Relationship Between Perceived Social Isolation and Medical vs. Recreational Marijuana Use Among Adults in the United States. Psychiatry Int (Basel). 2026;7(2):10.3390/psychiatryint7020055. doi:10.3390/psychiatryint7020055

54. SoftwareStataCorpLLC. Stata. 2026. URL: https://www.stata.com/company/ [accessed 2026-06-06]

55. Deary IJ, Corley J, Gow AJ, et al. Age-associated cognitive decline. Br Med Bull. 2009;92:135–152. doi:10.1093/bmb/ldp033

56. Karlamangla A, Tinetti M, Guralnik J, et al. Comorbidity in older adults: nosology of impairment, diseases, and conditions. J Gerontol A Biol Sci Med Sci. 2007;62(3):296–300. doi:10.1093/gerona/62.3.296

57. Andersen FK, Christensen K, Frederiksen H. Self-rated health and age: a cross-sectional and longitudinal study of 11,000 Danes aged 45-102. Scand J Public Health. 2007;35(2):164–171. doi:10.1080/14034940600975674

58. Offermann J, Wilkowska W, Laurentius T, et al. How age and health status impact attitudes towards aging and technologies in care: a quantitative analysis. BMC Geriatr. 2024;24(1):9. doi:10.1186/s12877-023-04616-4

59. Han Y, Zhang J, Qing W, et al. Influencing factors of digital health technology anxiety in the elderly: a systematic review and meta-analysis. Front Psychol. 2025;16:1645753. doi:10.3389/fpsyg.2025.1645753

60. Jason K, Carr D, Chen Z. Race-Ethnic Differences in the Effects of COVID-19 on the Work, Stress, and Financial Outcomes of Older Adults. J Aging Health. 2023;35(9):749–760. doi:10.1177/08982643231159705

61. Hollimon LA, Taylor KV, Fiegenbaum R, et al. Redefining and solving the digital divide and exclusion to improve healthcare: going beyond access to include availability, adequacy, acceptability, and affordability. Front Digit Health. 2025;7:1508686. doi:10.3389/fdgth.2025.1508686

62. Seligman ME. Learned helplessness. Annu Rev Med. 1972;23:407–412. doi:10.1146/annurev.me.23.020172.002203

63. Mannheim I, Weiss D, van Zaalen Y, et al. An “ultimate partnership”: Older persons’ perspectives on age-stereotypes and intergenerational interaction in co-designing digital technologies. Arch Gerontol Geriatr. 2023;113:105050. doi:10.1016/j.archger.2023.105050

64. Sweller J. Cognitive load during problem solving: Effects on learning. Cognitive science. 1988;12(2):257–85. doi:10.1207/s15516709cog1202_4

65. Fox JR, Park B, Lang A. When available resources become negative resources— The effects of cognitive overload on memory sensitivity and criterion bias. Commun Res 2007;34:277–296. doi:10.1177/0093650207300429

66. Jiang D, Kalyuga S, Sweller J. Comparing face-to-face and computer-mediated collaboration when teaching EFL writing skills. Educational Psychology. 2021;41(1):5–24. doi:10.1080/01443410.2020.1785399

67. Schakel W, Bode C, van der Aa HPA, et al. Exploring the patient perspective of fatigue in adults with visual impairment: a qualitative study. BMJ Open. 2017;7(8):e015023. doi:10.1136/bmjopen-2016-015023

68. Schakel W, Bode C, Elsman EBM, et al. The association between visual impairment and fatigue: a systematic review and meta-analysis of observational studies. Ophthalmic Physiol Opt. 2019;39(6):399–413. doi:10.1111/opo.12647

69. Veldman MHJ, Adanç B, van Rens GHMB, et al. Exploring cognitive overload in adults with visual impairment: The association between concentration and fatigue. Optom Vis Sci. 2024;101(11):646–651. doi:10.1097/OPX.0000000000002192

70. Humes LE, Young LA. Sensory-Cognitive Interactions in Older Adults. Ear Hear. 2016;37 Suppl 1(Suppl 1):52S–61S. doi:10.1097/AUD.0000000000000303

71. Brod C. Technostress: The human cost of the computer revolution. Cham: Springer. 1984.

72. Tarafdar M, Tu Q, Ragu-Nathan BS, Ragu-Nathan TS. The impact of technostress on role stress and productivity. Journal of Management Information Systems. 2007;24:301–328. doi:10.2753/MIS0742-1222240109

73. Kim SY, Park H, Kim H, et al. Technostress causes cognitive overload in high-stress people: Eye tracking analysis in a virtual kiosk test. Inf Process Manag. 2022;59(6):103093. doi:10.1016/j.ipm.2022.103093

74. Campbell FAK. Exploring internalized ableism using critical race theory. Disability & Society. 2008;23(2):151–162. doi:10.1080/09687590701841190

75. Campbell, F. K. (2009). Contours of ableism: The production of disability and abledness. Palgrave Macmillan UK. doi:10.1057/9780230245181

76. Butler RN. Age-ism: another form of bigotry. Gerontologist. 1969;9(4):243–246. doi:10.1093/geront/9.4_part_1.243

77. Allen JO. Ageism as a risk factor for chronic disease. Gerontologist. 2016;56(4):610–614. doi:10.1093/geront/gnu158

78. Allen JO, Solway E, Kirch M, et al. Experiences of Everyday Ageism and the Health of Older US Adults. JAMA Netw Open. 2022;5(6):e2217240. doi:10.1001/jamanetworkopen.2022.17240

79. Gudynaite G, Zamalijeva O, Pakalniskiene V, et al. Experiences of ageism and digital technology use among older adults. Front Psychol. 2025;16:1669321. doi:10.3389/fpsyg.2025.1669321

80. Choi EY, Kim Y, Chipalo E, et al. Does Perceived Ageism Widen the Digital Divide? And Does It Vary by Gender?. Gerontologist. 2020;60(7):1213–1223. doi:10.1093/geront/gnaa066

81. Livingston JD, Boyd JE. Correlates and consequences of internalized stigma for people living with mental illness: a systematic review and meta-analysis. Soc Sci Med. 2010;71(12):2150–2161. doi:10.1016/j.socscimed.2010.09.030

82. Mateus KS, Oliveira JS, Do Bú E, et al. Internalized ageism and late-life sexual health: gendered sexual meanings and a self-esteem-anxiety pathway to sexual dysfunction/distress. Aging Ment Health. 2026. doi:10.1080/13607863.2026.2656228

83. Fang Z, Liu Y, Peng B. Empowering older adults: bridging the digital divide in online health information seeking. Humanit Soc Sci Commun. 2024;11(1):1748. doi:10.1057/s41599-024-04312-7

84. Xin Y, Minyang L. Digital health literacy and self-efficacy among older adults: Mediating roles of social support and life satisfaction and the moderating role of health consciousness. Digit Health. 2026;12:20552076261415930. 2026. doi:10.1177/20552076261415930

