## Supplementary material for "Digital self-efficacy as a potential intermediary between vision impairment and daily internet use among older adults: A cross-sectional analysis of HINTS 2024": Multimedia Appendix

**Multimedia Appendix 1. Study flow chart of participants selection from HINTS 2024**

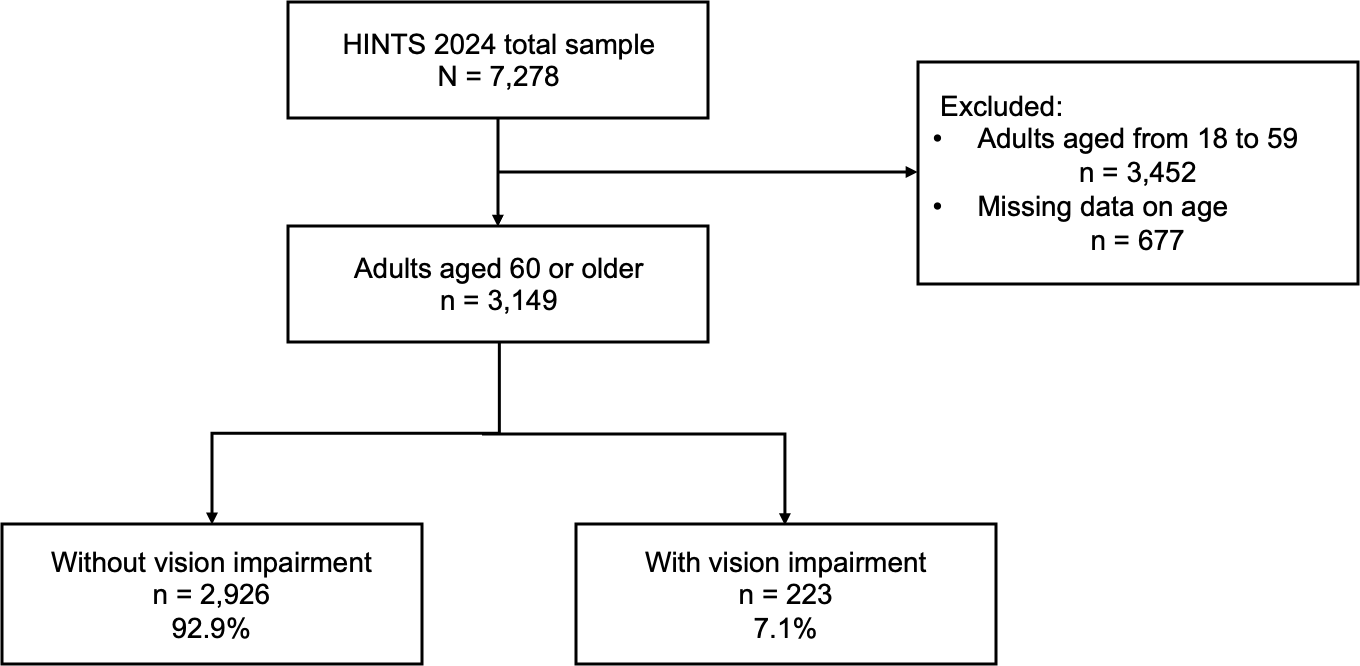

**Multimedia Appendix 2. The Strengthening the Reporting of Observational Studies in Epidemiology (STROBE) checklist**

STROBE Statement—Checklist of items that should be included in reports of ***cross-sectional studies***

|  | Item No | Recommendation | Location in a Manuscript |
| --- | --- | --- | --- |
| **Title and abstract** | 1 | (*a*) Indicate the study’s design with a commonly used term in the title or the abstract | Title, Abstract |
|  |  | (*b*) Provide in the abstract an informative and balanced summary of what was done and what was found | Abstract |
| Introduction | | |  |
| Background/rationale | 2 | Explain the scientific background and rationale for the investigation being reported | Introduction (Paragraph 1-4) |
| Objectives | 3 | State specific objectives, including any prespecified hypotheses | Introduction (Paragraph 5) |
| Methods | | |  |
| Study design | 4 | Present key elements of study design early in the paper | Methods (Study Design and Sample) |
| Setting | 5 | Describe the setting, locations, and relevant dates, including periods of recruitment, exposure, follow-up, and data collection | Methods (Study Design and Sample) |
| Participants | 6 | (*a*) Give the eligibility criteria, and the sources and methods of selection of participants | Methods (Study Design and Sample) |
| Variables | 7 | Clearly define all outcomes, exposures, predictors, potential confounders, and effect modifiers. Give diagnostic criteria, if applicable | Methods (Measures) |
| Data sources/ measurement | 8* | For each variable of interest, give sources of data and details of methods of assessment (measurement). Describe comparability of assessment methods if there is more than one group | Methods (Measures) |
| Bias | 9 | Describe any efforts to address potential sources of bias | Methods (Study Design and Sample, Statistical Analysis) |
| Study size | 10 | Explain how the study size was arrived at | Not reported |
| Quantitative variables | 11 | Explain how quantitative variables were handled in the analyses. If applicable, describe which groupings were chosen and why | Methods (Statistical Analysis) |
| Statistical methods | 12 | (*a*) Describe all statistical methods, including those used to control for confounding | Methods (Statistical Analysis) |
|  |  | (*b*) Describe any methods used to examine subgroups and interactions | Not applicable |
|  |  | (*c*) Explain how missing data were addressed | Methods (Statistical Analysis) |
|  |  | (*d*) If applicable, describe analytical methods taking account of sampling strategy | Methods (Statistical Analysis) |
|  |  | (*e*) Describe any sensitivity analyses | Methods (Statistical Analysis) |
| Results | | |  |
| Participants | 13* | (a) Report numbers of individuals at each stage of study—eg numbers potentially eligible, examined for eligibility, confirmed eligible, included in the study, completing follow-up, and analysed | Methods (Study Design and Sample, Multimedia Appendix 1),  Results (Sample Characteristics) |
|  |  | (b) Give reasons for non-participation at each stage | Methods (Study Design and Sample, Multimedia Appendix 1) |
|  |  | (c) Consider use of a flow diagram | Methods (Study Design and Sample, Multimedia Appendix 1) |
| Descriptive data | 14* | (a) Give characteristics of study participants (eg demographic, clinical, social) and information on exposures and potential confounders | Results (Sample Characteristics, Table 1) |
|  |  | (b) Indicate number of participants with missing data for each variable of interest | Results (Table 1-2) |
| Outcome data | 15* | Report numbers of outcome events or summary measures | Results (Survey-weighted Prevalences of Digital Access, Skills, Self-efficacy, and Frustration, Table 2) |
| Main results | 16 | (*a*) Give unadjusted estimates and, if applicable, confounder-adjusted estimates and their precision (eg, 95% confidence interval). Make clear which confounders were adjusted for and why they were included | Results (Multivariable Logistic Regression Analyses, Table 3-6) |
|  |  | (*b*) Report category boundaries when continuous variables were categorized | Not applicable |
|  |  | (*c*) If relevant, consider translating estimates of relative risk into absolute risk for a meaningful time period | Results (Multivariable Logistic Regression Analyses, Table 5) |
| Other analyses | 17 | Report other analyses done—eg analyses of subgroups and interactions, and sensitivity analyses | Results (Multivariable logistic regression analyses, Multimedia Appendix 3) |
| Discussion | | |  |
| Key results | 18 | Summarise key results with reference to study objectives | Discussion (Principal findings (Paragraph 1)) |
| Limitations | 19 | Discuss limitations of the study, taking into account sources of potential bias or imprecision. Discuss both direction and magnitude of any potential bias | Discussion (Strengths and Limitations) |
| Interpretation | 20 | Give a cautious overall interpretation of results considering objectives, limitations, multiplicity of analyses, results from similar studies, and other relevant evidence | Discussion (Principal findings (Paragraph 1-9), Clinical Implications, Strengths and Limitations) |
| Generalisability | 21 | Discuss the generalisability (external validity) of the study results | Discussion (Strengths and Limitations) |
| Other information | | |  |
| Funding | 22 | Give the source of funding and the role of the funders for the present study and, if applicable, for the original study on which the present article is based | Acknowledgments |

*Give information separately for exposed and unexposed groups.

**Note:** An Explanation and Elaboration article discusses each checklist item and gives methodological background and published examples of transparent reporting. The STROBE checklist is best used in conjunction with this article (freely available on the Web sites of PLoS Medicine at http://www.plosmedicine.org/, Annals of Internal Medicine at http://www.annals.org/, and Epidemiology at http://www.epidem.com/). Information on the STROBE Initiative is available at www.strobe-statement.org.

**Multimedia Appendix 3. Sensitivity analyses: Survey-weighted multivariable logistic regression: Association between vision impairment and digital access, skills, self-efficacy, and frustration among older adults,** **adjusting for income instead of education (n = 2,848)^a-c^**

| Outcomes | Unadjusted OR (95% CI) | *P* value | Adjusted OR (95% CI) | *P* value | Adjusted predicted marginal proportions (95% CI) | Risk difference (95% CI) |
| --- | --- | --- | --- | --- | --- | --- |
| **Digital access** |  |  |  |  |  |  |
| Daily internet use^c^ | 0.45 (0.26 to 0.79) | .007 | 0.62 (0.37 to 1.06) | .078 | 0.76 (0.69 to 0.83) | -0.06 (-0.14 to 0.01) |
| Smartphone use | 0.71 (0.36 to 1.40) | .314 | 0.89 (0.37 to 2.17) | .802 | 0.85 (0.75 to 0.95) | -0.01 (-0.11 to 0.08) |
| Computer use | 0.49 (0.29 to 0.83) | .010 | 0.64 (0.36 to 1.16) | .137 | 0.67 (0.56 to 0.78) | -0.08 (-0.19 to 0.03) |
| Tablet use | 0.57 (0.30 to 1.08) | .085 | 0.71 (0.36 to 1.39) | .314 | 0.39 (0.24 to 0.55) | -0.08 (-0.24 to 0.08) |
| Wearable device use | 0.81 (0.34 to 1.89) | .613 | 1.22 (0.51 to 2.91) | .650 | 0.24 (0.09 to 0.38) | 0.03 (-0.11 to 0.18) |
| **Digital skills** |  |  |  |  |  |  |
| Used the internet to look for health information^d^ | 0.88 (0.44 to 1.78) | .722 | 0.72 (0.31 to 1.67) | .437 | 0.76 (0.62 to 0.90) | -0.05 (-0.20 to 0.09) |
| Used the internet to send a message to a healthcare provider^d^ | 0.84 (0.49 to 1.46) | .533 | 0.91 (0.45 to 1.84) | .799 | 0.62 (0.46 to 0.78) | -0.02 (-0.18 to 0.14) |
| Used the internet to view medical test results^d^ | 0.85 (0.49 to 1.49) | .572 | 0.76 (0.37 to 1.54) | .434 | 0.67 (0.53 to 0.82) | -0.06 (-0.20 to 0.09) |
| Used the internet to make an appointment with a healthcare provider^d^ | 0.66 (0.37 to 1.17) | .153 | 0.66 (0.33 to 1.32) | .236 | 0.47 (0.30 to 0.63) | -0.10 (-0.26 to 0.07) |
| Used telehealth to receive care from healthcare professional^e^ | 1.21 (0.67 to 2.18) | .525 | 1.01 (0.84 to 1.77) | .966 | 0.30 (0.27 to 0.34) | 0.00 (-0.14 to 0.14) |
| **Digital self-efficacy** |  |  |  |  |  |  |
| Confidence in using apps without help^f^ | 0.49 (0.31 to 0.80) | .005 | 0.50 (0.29 to 0.88) | .017 | 0.50 (0.37 to 0.63) | -0.15 (-0.28 to -0.02) |
| **Digital frustration** |  |  |  |  |  |  |
| Frustration to learn how to use new technology^g^ | 2.31 (1.07 to 5.00) | .034 | 2.02 (0.91 to 4.50) | .082 | 0.79 (0.66 to 0.92) | 0.13 (0.01 to 0.26) |

Abbreviation: CI: confidence interval, OR: odds ratio

^a^ All models used the jackknife replication method to ensure the sample weighting.

^b^ All models were adjusted for age, sex, race/ethnicity, income, and the number of comorbidities.

^c^ Daily internet use: Yes = 1 (responded ‘More than once per day’ or ‘About once per day’), No = 0 (responded ‘A few times a week,’ ‘Less than once per week,’ ‘Rarely,’ or ‘Never’)

^d^ Respondents who answered ‘Never’ to the question on frequency of internet use were treated as missing data and excluded in the analyses for the following four variables: looking for health information, sending a message to a healthcare provider, viewing medical test results, and making an appointment with a healthcare provider.

^e^ Used telehealth to receive care from healthcare professional: Yes = 1 (responded ‘Yes, by video,’ ‘Yes, by phone call (voice only with no video),’ or ‘Yes, some by video and some by phone call.’), No = 0 (responded ‘No telehealth visits in the past 12 months’)

^f^ Confidence in using apps without help: Yes = 1 (responded ‘strongly agree’ or ‘somewhat agree’), No = 0 (responded ‘somewhat disagree’ or ‘strongly disagree’)

^g^ Frustration to learn how to use new technology: Yes = 1 (responded ‘strongly agree’ or ‘somewhat agree’), No = 0 (responded ‘somewhat disagree’ or ‘strongly disagree’)
